# Two anti-phase spatial modes and a candidate spatial-persistence regime transition of SARS-CoV-2 in Japan: a 159-week prefecture-level sentinel surveillance study

**DOI:** 10.64898/2026.05.24.26353972

**Authors:** Takashi Nakano, Daisuke Onozuka, Yoichi Ikeda, Kouhei Washiyama, Yoshihiro Takashima

**Affiliations:** Research Center for Nuclear Physics, The University of Osaka, Osaka, Japan; Center for Infectious Disease Education and Research (CiDER), The University of Osaka, Osaka, Japan; Center for Advanced Modalities and DDS (CAMaD), The University of Osaka, Osaka, Japan; Department of Post-Infectious Diseases Therapeutics, Graduate School of Medicine, The University of Osaka, Osaka, Japan

**Author notes:** **Corresponding author:** Takashi Nakano, Research Center for Nuclear Physics, The University of Osaka.

**Keywords:** COVID-19, SARS-CoV-2, spatial epidemiology, sentinel surveillance, Moran’s I, Shannon entropy, change-point detection, metapopulation dynamics, Japan, regime transition, endemic transition

## Abstract

**Background:** On 8 May 2023 the Japanese Ministry of Health, Labour and Welfare reclassified COVID-19 under the Infectious Disease Control Law from a designated infectious disease (with case-by-case reporting requirements comparable to those of a Category-2 disease) to a Category-5 (“Class-5”) notifiable disease, joining the same category as seasonal influenza and most other endemic respiratory infections. Under this regime, COVID-19 case counts are reported weekly from a nationwide network of sentinel medical facilities (initially approximately 5,000, reduced to approximately 3,000 following an April 2025 surveillance reform), and individual case reporting is no longer required. We aimed to characterize the spatial topology of COVID-19 epidemics under this sentinel-surveillance regime and to detect, in a data-driven manner, any structural change in epidemic dynamics over this period.

**Methods:** We analyzed weekly per-sentinel-facility COVID-19 case counts in all 47 prefectures of Japan from 2023-W17 to 2026-W19 (159 weeks). For each week we computed the Shannon pseudo-entropy *S* of the prefecture-share distribution and global, local, and time-lagged Moran’s *I* across a 92-edge contiguity-based adjacency matrix. To identify any structural change in a data-driven manner, we adopted a two-stage approach motivated by an empirical regularity established in §3: we first verified the wave-amplitude-invariant *entropy ceiling* (*S*_max ≥ 3.80 in all five pre-transition waves), then restricted change-point detection to the weeks after *S(t)* last attained this ceiling, applying PELT, CUSUM, and Bai–Perron sup-F within this restricted region. Seasonal structure was characterized by truncated Fourier regression with first-order autoregressive errors (Cochrane–Orcutt) over harmonic orders *K* ∈ {1, …, 6}; between-period comparisons used moving block bootstrap as the principal inferential statistic.

**Results:** The five epidemic waves during 2023–2025 followed a stereotyped spatial template in which *S(t)* traced a characteristic U-shape around each peak, with a wave-amplitude-invariant entropy ceiling reaching on average 99.4% of the theoretical maximum ln 47 (range 3.820– 3.836, SD 0.006). The last week in which *S(t)* attained this entropy ceiling was 2025-W42.

Restricting change-point detection to the 29 subsequent weeks, PELT and CUSUM localised the structural break to late 2025: PELT identified 2025-W48 (robust across penalty values ≥ σ^2^·ln(*n*) and across entropy-ceiling thresholds 3.78–3.82) and CUSUM peaked at 2025-W50 (*p* < 0.0001), placing the break within a two-week window centred on late November 2025. Bai– Perron sup-F peaked later at 2026-W02 (*p* = 0.062, with reduced power on *n* = 29). We adopted 2025-W48 as the principal change-point, defining 135 pre-transition weeks and 24 post-transition weeks. Two anti-phase spatial modes were identified in the pre-transition record: a summer-onset Okinawa-seeded Kyushu cascade (Mode A; annual peak epi week 26) and a winter-onset Tohoku-centred connected-cluster mode (Mode B; annual peak epi week 51), approximately 25 epi weeks out of phase. After the regime transition, this ceiling was not attained, and the spatial-persistence ratio *I*(τ = 8 wk)/*I*(0) shifted from a highly variable distribution centred near 0.27 (pre-transition, 125 weeks) to a tightly clustered distribution around 0.89 (post-transition, 24 weeks); the mean difference was 0.62 (95% bootstrap CI 0.32 to 0.90; moving block bootstrap *p* < 0.0001 across block lengths 1–12). The principal finding remained significant under autoregressive-augmented null models and was robust to adjacency-matrix choice, the April 2025 surveillance reform, harmonic order *K* ∈ {1, …, 6}, and Okinawa exclusion.

**Conclusions:** Data-driven analysis of 159 weeks of Japanese sentinel surveillance identifies a candidate spatial-persistence regime transition emerging in late November 2025, in which the spatial structure of weekly case shares persists for at least 8 weeks rather than dissipating as in pre-transition. The transition coincides with loss of the wave-amplitude-invariant entropy ceiling and with absence of the Mode A signature through the observed post-transition period. The recent uptick in Okinawa case shares (continuing through 2026-W19) leaves open whether the Mode A signature is structurally suppressed or merely deferred; observation through summer 2026 is required to distinguish a sustained shift from a transient anomaly.

## 1. Introduction

More than five years after the emergence of SARS-CoV-2, most national surveillance systems have transitioned from universal case ascertainment to sentinel-based monitoring as the public-health priority shifted from containment to trend characterization. In Japan, this transition occurred on 8 May 2023 with the reclassification of COVID-19 from a designated infectious disease (with case-by-case reporting comparable to Category-2 control measures) to a Category-5 (Class-5) notifiable disease under the Infectious Disease Control Law [2, 3]. Since this reclassification, weekly counts of new cases per sentinel medical facility have been reported by all 47 prefectures, providing the first opportunity to characterize SARS-CoV-2 dynamics in Japan under a stable post-pandemic surveillance regime. The accumulating record now spans nearly three years and approximately five distinguishable epidemic waves, raising the question of whether the post-Class-5 epidemiology of COVID-19 has reached a stable endemic state or is still undergoing structural change.

Existing analyses of post-Class-5 COVID-19 in Japan have focused predominantly on the temporal structure of nationally aggregated case counts: wave amplitudes, peak-to-peak intervals, age stratification, and the agreement between sentinel and other indicators such as school-absentee surveillance [4, 5]. The spatial structure of post-Class-5 epidemics has received less attention, although prior work using prefecture-level data from the universal-reporting era identified substantial geographic heterogeneity, with Okinawa typically exhibiting the highest mean reproduction number and Hokkaido the lowest [5]. Our group has previously developed complementary approaches to COVID-19 spread quantification, including the K-value [6], the broken-link compartment model [7], and Theil-index decomposition of regional disproportionality [8]. The current study extends this line of work into prefecture-level spatial topology by combining an information-theoretic concentration measure with spatial-autocorrelation statistics. Two methodological frameworks have been independently established for quantifying spatial structure in surveillance data: information-theoretic concentration indices (Theil index, Shannon entropy) [8, 9], which summarize the degree to which cases are unevenly distributed across geographic units, and spatial autocorrelation statistics (Moran’s I, local indicators of spatial association) [10–13], which characterize the degree to which high-incidence units are geographically clustered. The two frameworks address conceptually distinct questions: a single-number summary of concentration cannot, in principle, distinguish between cases concentrated in geographically connected regions versus cases concentrated in scattered isolated locations. Combining the two approaches has been advocated in spatial-epidemiology methodology [12, 14] but has not been systematically applied, to our knowledge, to Japanese COVID-19 surveillance data.

In this study we apply both frameworks — Shannon pseudo-entropy and global, local, and time-lagged Moran’s I — to weekly per-sentinel-facility COVID-19 case counts in all 47 prefectures of Japan over a 159-week period spanning the entire Class-5 sentinel-surveillance period covered by this study (2023-W17 to 2026-W19; the Class-5 designation remains in effect at the time of writing). We hypothesized, on the basis of Wagatsuma et al.’s [5] regional reproduction-number heterogeneity and the metapopulation framework established for measles by Grenfell, Bjørnstad, and Kappey [15–17], that nationwide outbreaks in Japan would exhibit identifiable spatial-mode structure that varies seasonally. The combination of pseudo-entropy and Moran’s I provides a principled means of testing this hypothesis: spatial modes that are invisible to either statistic in isolation should become evident when the two are examined jointly. We further sought to characterize whether the spatial structure has remained stable across the post-Class-5 period or has undergone identifiable structural change. A key methodological commitment of the present study is that any such structural change should be identified in a *data-driven* manner — that is, by formal change-point detection applied to the full 159-week series — rather than by pre-specifying any anomalous period in advance. The questions addressed are therefore: (i) does the post-Class-5 SARS-CoV-2 epidemic in Japan exhibit a stable wave-template spatial structure under the assumption of stationarity? (ii) within this template, are there qualitatively distinct seasonal modes of spatial spread? and (iii) when the assumption of stationarity is relaxed, do the data themselves identify a structural change, and if so, what is the nature of the post-transition behaviour?

## 2. Methods

### Study design

This is an ecological time-series study using nationally aggregated, prefecture-level COVID-19 surveillance data published by the Japanese Ministry of Health, Labour and Welfare (MHLW). The unit of analysis is the prefecture-week, with all 47 prefectures of Japan observed for 159 consecutive weeks. The study was based entirely on aggregate, publicly available data with no patient-level or identifying information; consequently, ethical review was not required.

### Setting and data sources

Following the reclassification of COVID-19 from a designated infectious disease (with case-by-case reporting comparable to Category-2 control measures) to a Category-5 (Class-5) notifiable disease under Japan’s Infectious Disease Control Law on 8 May 2023 [2], universal case reporting was discontinued and replaced by a sentinel surveillance reporting system [3]. Under this system, weekly COVID-19 case counts were reported by a nationwide network of approximately 5,000 designated sentinel medical facilities (the pediatric and internal-medicine sentinel sites) across the 47 prefectures, which the MHLW aggregates and publishes online.

We conducted an ecological study using aggregated weekly prefecture-level data. We obtained weekly per-sentinel-facility case counts of COVID-19, stratified by all 47 prefectures, from the MHLW publicly available reports at https://www.mhlw.go.jp/stf/seisakunitsuite/bunya/0000121431_00086.html. The dataset spans 159 consecutive weeks from 2023-W17 (week of 24 April 2023, immediately preceding the 8 May 2023 reclassification) to 2026-W19. Each week runs Monday through Sunday and is referenced throughout the manuscript by its Japanese epidemiological-week label (epi week, indexed 1–52 or 1–53 within each calendar year) and the date of its mid-point Thursday; harmonic regressions use the epi week index of each surveillance week as the continuous time variable. The first two weeks of the series (2023-W17 to 2023-W18) were collected under the pre-reclassification universal-reporting regime; MHLW retrospectively reports prefecture-level sentinel case counts for these weeks, and we include them in the principal analysis. We use the data as published, without further smoothing or imputation.

On 7 April 2025 (epi week 15), the sentinel surveillance system underwent a structural reform with the launch of the integrated Acute Respiratory Infection (ARI) surveillance program, which altered the criteria for sentinel facility designation and the composition of the reporting facility pool, reducing the national sentinel network to approximately 3,000 designated facilities. We address the implications of this reform through dedicated sensitivity analyses (Supplementary Materials).

### Outcome measures: Pseudo-entropy and Kullback–Leibler decomposition

Let *x*_i_*(t)* denote the per-sentinel-facility weekly case count for prefecture *i* (i = 1, …, 47) at week *t*. We define the prefecture-share probability

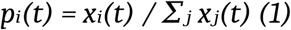

which by construction satisfies Σ_i_ *p*_i_(t) = 1 for all *t*. Because Japan’s sentinel surveillance system is calibrated such that designated medical facilities are distributed broadly in proportion to prefectural healthcare-utilization patterns [3], the per-sentinel-facility report rate *x*_i_ serves as a national-comparable proxy for prefectural-level disease intensity. The share variable *p*_i_*(t)* therefore captures the relative spatial distribution of disease intensity across prefectures, removing the absolute incidence scale. The same construction was used in our prior work [8], where it was applied to U.S. state-level case counts as input to the Theil index of regional disproportionality.

As the principal indicator of nationwide epidemic spatial uniformity, we computed the Shannon entropy of the prefecture-share distribution:

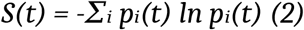

Hereafter we refer to *S(t)* as the pseudo-entropy of the spatial case distribution. The quantity is bounded between 0 (all reports concentrated in a single prefecture) and ln 47 ≈ 3.850 (uniform per-sentinel-facility report rate across all prefectures); higher values therefore indicate spatial uniformity, lower values indicate spatial concentration. The maximum of *S(t)* attained during each inter-wave plateau — termed the *entropy ceiling* of the wave template — is a wave-amplitude-invariant feature that we characterise empirically in §3.

The Kullback–Leibler divergence of *p(t)* from the uniform distribution **u** = (1/47, …, 1/47) is

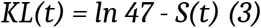

which we further decomposed into regional contributions by partitioning the 47 prefectures into eight standard regional blocks (Hokkaido [1]; Tohoku [6]; Kanto [7]; Chubu [9]; Kinki [7]; Chugoku [5]; Shikoku [4]; Kyushu+Okinawa [8]). The contribution of region R to the divergence is

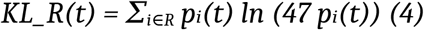

with Σ_R_ KL_R = KL. This decomposition is sign-informative: in aggregate, a region whose report rates typically exceed the national mean contributes positively, whereas a region falling below the mean contributes negatively.

### Outcome measures: Global, local, and time-lagged Moran’s I

To distinguish between spatially clustered and spatially scattered concentration patterns at the same level of *S*, we computed global Moran’s I [10, 11] using a contiguity-based spatial weights matrix **W** (47 × 47, symmetric, zero diagonal). Two prefectures *i* and *j* were assigned weight *W*_ij_ = 1 if they shared a land border or were connected by a major fixed link (Seikan Tunnel between Hokkaido and Aomori; Kanmon Bridge/Tunnel between Yamaguchi and Fukuoka; Seto Ohashi Bridge between Okayama and Kagawa; Shimanami Kaido between Hiroshima and Ehime; Akashi Kaikyo and Onaruto Bridges between Hyogo and Tokushima; the Amami island chain serving as a stepping-stone between Kagoshima and Okinawa). All other entries were zero. The resulting **W** contained 92 non-zero edges. Robustness to alternative specifications of **W** is examined in the Supplementary Materials.

For each week, global Moran’s I was computed as

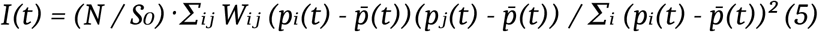

where *N* = 47, *S*_0_ = Σ_ij_ *W*_ij_ = 184 (twice the number of edges), and 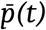 is the mean of *p*_i_*(t)* across prefectures (= 1/47). Statistical significance of each weekly *I(t)* was assessed by 9,999 random permutations of the prefecture labels [12]; the pseudo-p-value was the rank of the observed *I(t)* within the permutation distribution. These within-week permutation tests assess spatial homogeneity conditional on the weekly case total; the temporal autocorrelation of the resulting *I(t)* time series is treated separately in the between-period analyses described below.

To quantify the temporal persistence of spatial structure, we computed time-lagged bivariate Moran’s I at lags τ ∈ {0, 1, 2, 4, 8} weeks:

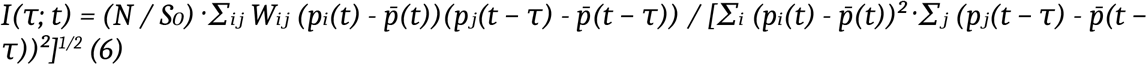

Positive *I(*τ) indicates that high prefecture-share neighbours τ weeks ago predict high prefecture-share values now. The standardized ratio *I(*τ*)/I(0)* — termed hereafter the *spatial-persistence ratio* at lag τ — provides a scale-free measure of the temporal persistence of spatial structure: values close to 1 indicate that the spatial pattern is preserved over τ weeks, values close to 0 indicate that the pattern has dissolved or migrated. This ratio is the principal quantity used to characterise the regime transition in §3.

### Harmonic regression with autoregressive errors

Seasonal structure in *S(t), I(t)*, and the regional KL contributions was characterised by truncated Fourier regression of order *K*:

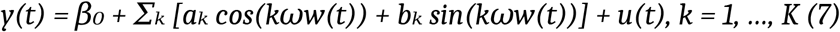

where *w(t)* is the fractional epi-week index of the mid-point Thursday of week *t* (taking values in the range 1 ≤ *w* ≤ 53), ω = 2π/52.1775 rad week^−1^ (so that ω·52.1775 = 2π corresponds to one solar year), and the residual process *u(t)* is modeled as first-order autoregressive,

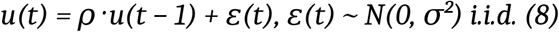

Equations (7)–(8) were estimated jointly by iterative Cochrane–Orcutt (CO) regression to convergence. Throughout, a circumflex denotes a sample estimate obtained from this fit (e.g., 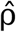 estimates the autoregressive coefficient ρ, and 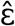 the model innovations ε). The AR(1) augmentation accommodates the strong residual serial dependence 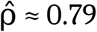 for *S(t)* pre-transition residuals at *K* = 2) that would otherwise invalidate independence-based inference, and corresponds to a discretised Ornstein–Uhlenbeck noise process with characteristic correlation time 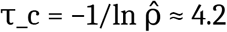. Goodness of fit is reported as the AR(1)-corrected R^2^:

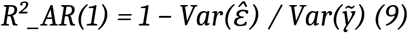

where 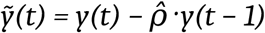 is the quasi-differenced response. This statistic is interpretable as the proportion of innovation variance explained by the deterministic harmonic component, and is generally substantially smaller than the corresponding OLS R^2^ because the AR(1) process accounts for much of the apparent seasonal variation through its persistence alone.

The choice of harmonic order *K* was examined empirically across *K* ∈ {1, 2, 3, 4, 5, 6} using both AIC and BIC computed on the AR(1)-augmented likelihood. Results are reported in §3 and the Supplementary Materials.

### Change-point detection

To identify any structural change in the spatial-statistical time series without pre-specifying any anomalous period, we adopted a two-stage data-driven approach motivated by the empirical regularity established in §3 (the wave-amplitude-invariant entropy ceiling).

#### Stage 1 (establishment of empirical regularity)

From the pre-transition data we verified that the inter-wave plateau maxima of *S(t)* satisfy *S*_max ≥ 3.80 in every pre-transition wave (mean 3.827, SD 0.006 across five waves; Table 1). This regularity defines an *entropy-ceiling regime* in which *S(t)* recurrently revisits values close to its theoretical upper bound ln 47 ≈ 3.850.

**Table 1.** Wave amplitude versus inter-wave entropy ceiling, five pre-transition waves.

| Wave | Peak epi week | Peak amp. (per sentinel) | $S_{\max}$ epi week | $S_{\max}$ | % of $\ln 47$ |
| --- | --- | --- | --- | --- | --- |
| Wave 1 | 2023-W35 | 20.7 | 2023-W38 | 3.836 | 99.6% |
| Wave 2 | 2024-W05 | 16.6 | 2024-W08 | 3.827 | 99.4% |
| Wave 3 | 2024-W30 | 15.6 | 2024-W32 | 3.827 | 99.4% |
| Wave 4 | 2025-W02 | 7.6 | 2025-W05 | 3.820 | 99.2% |
| Wave 5 | 2025-W34 | 9.8 | 2025-W41 | 3.826 | 99.4% |
| Pre-transition mean | — | 14.1 | — | 3.827 (SD 0.006) | 99.4% |
| Post-transition (n = 24) | — | — | 2026-W18 | 3.689 | 95.8% |

*Peak amplitude is the nationwide weekly per-sentinel-facility case count $((1/47)\sum_i x_i(t))$ at the wave peak. $S_{\max}$ is the local maximum of $S(t)$ in the $\pm 8$ -week window around the corresponding peak. The theoretical maximum attainable by $S$ is $\ln 47 \approx 3.850$ , achieved when prefecture-shares are exactly uniform. The narrow 0.43% relative spread of $S_{\max}$ across the five pre-transition waves stands in contrast to the 2.7-fold spread of peak amplitudes; the post-transition row is given for comparison.*

#### Stage 2 (restricted change-point detection)

We defined the operational onset of any regime transition as the last week in which *S(t)* attained the entropy ceiling, operationalised as *S(t)* ≥3.80. In the 159-week dataset, this was 2025-W42 (S = 3.820), corresponding to the post-peak plateau of the fifth pre-transition wave. We then restricted the change-point search to the subsequent 29 weeks (2025-W43 to 2026-W19) and applied three independent change-point detection methods within this restricted region:

1. **PELT (Pruned Exact Linear Time)** [18] with an L_2_ cost function, implemented via the Python `ruptures` library [19] (version 1.1). The PELT penalty was chosen as pen = 2σ^2^·ln(*n*) (BIC-style); sensitivity to penalty values in the range 0.5×–4× of this baseline is reported as a robustness check.
2. **CUSUM** [20], using the sup-CUSUM test statistic applied to the de-meaned series; the empirical *p*-value was obtained from 5,000 random permutations.
3. **Bai–Perron sup-F test** for a single structural break in the harmonic regression model of equation (7) [21]. We computed the Chow *F*-statistic at every candidate breakpoint within the central 80% of the restricted region (trimming π_0_ = 0.10), and obtained the empirical *p*-value by 1,000 wild-bootstrap replicates.
4. Sensitivity to the entropy-ceiling threshold choice was assessed across the range 3.78 ≤ threshold ≤ 3.82, all of which fall below the pre-transition wave-ceiling minimum of 3.820 (Table 1) and therefore exclude none of the five pre-transition wave plateaus from the entropy-ceiling regime. The threshold was chosen below the minimum observed pre-transition wave ceiling (3.820) and was used only to define the last return to the empirically established ceiling regime; the threshold itself is not used to optimise the detected change-point and is not a tunable hyperparameter of the principal analysis.

This two-stage approach achieves three methodological benefits over global change-point detection: it directly tests the entropy-ceiling regularity established in §3 rather than introducing a separate change-point hypothesis; it excludes from the search region the wave-related transient dips in *S(t)* (most notably the deep early-July 2025 single-week minimum) that would confound global change-point estimates; and it produces a directly interpretable change-point — the week from which the system departed from the established entropy-ceiling regime. The restricted analysis was not designed to search for an arbitrary break after visual inspection of the data, but to test whether the empirically recurrent entropy-ceiling regime failed after its final observed return; the search region is fixed by t_last rather than chosen post hoc.

For completeness, we also applied PELT, CUSUM, Bai-Perron sup-F, and Bayesian online change-point detection (BOCPD) [22] to the full 159-week series as a baseline analysis; results of this global analysis are reported in the Supplementary Materials. The principal change-point identified by the restricted analysis (Stage 2) was used to define a *pre-transition* period and a *post-transition* period for all subsequent between-period analyses, as detailed in §3.

### Statistical analysis

Between-period comparisons (pre-transition vs. post-transition; summer-window vs. winter-window) involve two samples of weekly time-series observations with substantial within-sample serial dependence (within-sample lag-1 autocorrelations in the range ρ_1_= 0.78–0.90 across the principal quantities *I(t), S(t)*, KL contributions, and time-lagged Moran’s I). The **moving block bootstrap** [23, 24] is our sole inferential statistic for between-period comparisons. For each comparison, 10,000 replicates were generated by independently resampling fixed-length blocks from each group; the observed difference in means was compared against the bootstrap distribution, and a two-sided *z*-statistic and *p*-value were computed using the bootstrap standard error. The block length was set to 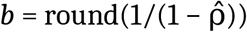 per group, with sensitivity across *b* ∈ {1, 2, 4, 6, 8, 10, 12} reported as a robustness check.

Confidence intervals for between-period mean differences were obtained from the 2.5th and 97.5th percentiles of the bootstrap distribution. The moving block bootstrap preserves the within-sample serial dependence that would invalidate any standard parametric comparison.

For continuity with prior reporting, we also report a one-sample sign test on the post-transition residuals from the pre-transition harmonic fit (which depends only on the marginal probability of a positive deviation and is therefore robust to residual serial dependence). The two-sided significance threshold is α = 0.05; bootstrap p-values below 0.0001 are reported as p < 0.0001.

### Sensitivity analyses

To assess the robustness of our principal findings against modelling choices and surveillance-system changes, we conducted five pre-specified sensitivity analyses: (i) alternative specifications of the spatial weights matrix **W** (six variants ranging from contiguity-only to continuous distance-decay; see Supplementary Materials); (ii) harmonic-order choice *K* ∈ {1, …, 6} with AR(1) errors; (iii) leave-one-prefecture-out analysis of the AIC ranking between K = 2 and K = 3 for *I(t)*, identifying prefectures whose non-sinusoidal sharp peaks drive any apparent K = 3 preference; (iv) sensitivity to the 7 April 2025 surveillance reform, by repeating the principal between-period comparison restricted to the post-reform period; and (v) Okinawa exclusion, with all spatial-statistical quantities recomputed on the 46-prefecture subset.

Detailed results of (i)–(v) are reported in the Supplementary Materials; the main text reports only the principal findings and selected indicators of robustness.

### Software and reproducibility

All analyses were performed in Python 3.11 (NumPy 1.26, SciPy 1.12, pandas 2.2, scikit-learn 1.4, matplotlib 3.8, statsmodels 0.14, ruptures 1.1). Random permutation tests for Moran’s I used a fixed random seed (42) for reproducibility. The complete analysis pipeline (data ingestion, computation of *S*, Moran’s I, harmonic regression, change-point detection, sensitivity analyses, and figure generation) is available at https://github.com/takashi7nakano/covid-spatial-entropy-japan (archived at Zenodo, DOI: 10.5281/zenodo.20359405). The processed prefecture-week dataset and adjacency matrix are deposited as Supplementary Data.

### Patient and public involvement

This study used aggregated public surveillance data without direct patient or public involvement in study design or interpretation. Findings will be communicated to the public through a press release accompanying publication and through medRxiv preprint deposit.

## 3. Results

### Sample characteristics

The dataset comprised 159 weeks of prefecture-level COVID-19 sentinel reports from 2023-W17 to 2026-W19 (24 April 2023 to 10 May 2026), covering all 47 prefectures of Japan (7,473 prefecture-weeks; no missing values). The pseudo-entropy *S(t)* ranged from 3.541 in 2025-W27 to 3.836 in 2023-W38, corresponding to an effective spatial occupancy *W*_eff = exp(*S*) ranging from 34.5 to 46.3 of the maximum 47 prefectures. Moran’s *I* ranged from −0.143 in 2025-W21 to 0.736 in 2024-W36, with 144/159 weeks (90.6%) showing positive spatial autocorrelation significant at the within-week permutation *p* < 0.05 level.

### The 2023–2025 wave template

During the 135 pre-transition weeks, Japan experienced five distinguishable epidemic waves with peak weeks at approximately 28 August 2023, 1 February 2024, 25 July 2024, 9 January 2025, and 21 August 2025. Across all five waves, *S(t)* exhibited a stereotyped trajectory: a local minimum approximately 7 weeks before each peak (mean lag 7.2 weeks, range 5–9), followed by ascent to a local maximum approximately 3 weeks after each peak (mean lag 3.4 weeks, range 2–5), and a subsequent inter-wave plateau of high *S* indicating spatial uniformity. The cross-correlation between *S(t)* and log nationwide cases (computed on pre-transition weeks) was maximized at lag +4 weeks (Pearson *r* = 0.675, 95% block-bootstrap CI 0.580–0.751), confirming that nationwide spatial uniformity follows rather than precedes peaks in incidence (Figure 1A). Although the absolute peak amplitudes of successive waves declined over the 2023–2025 period (from ∼21 to ∼10 reports per sentinel facility), the seasonal range of *S* expanded (Pearson *r* = −0.57, 95% block-bootstrap CI −0.78 to −0.21), indicating that the spatial concentration of activity at troughs intensified as overall epidemic intensity declined.

**Figure 1.**
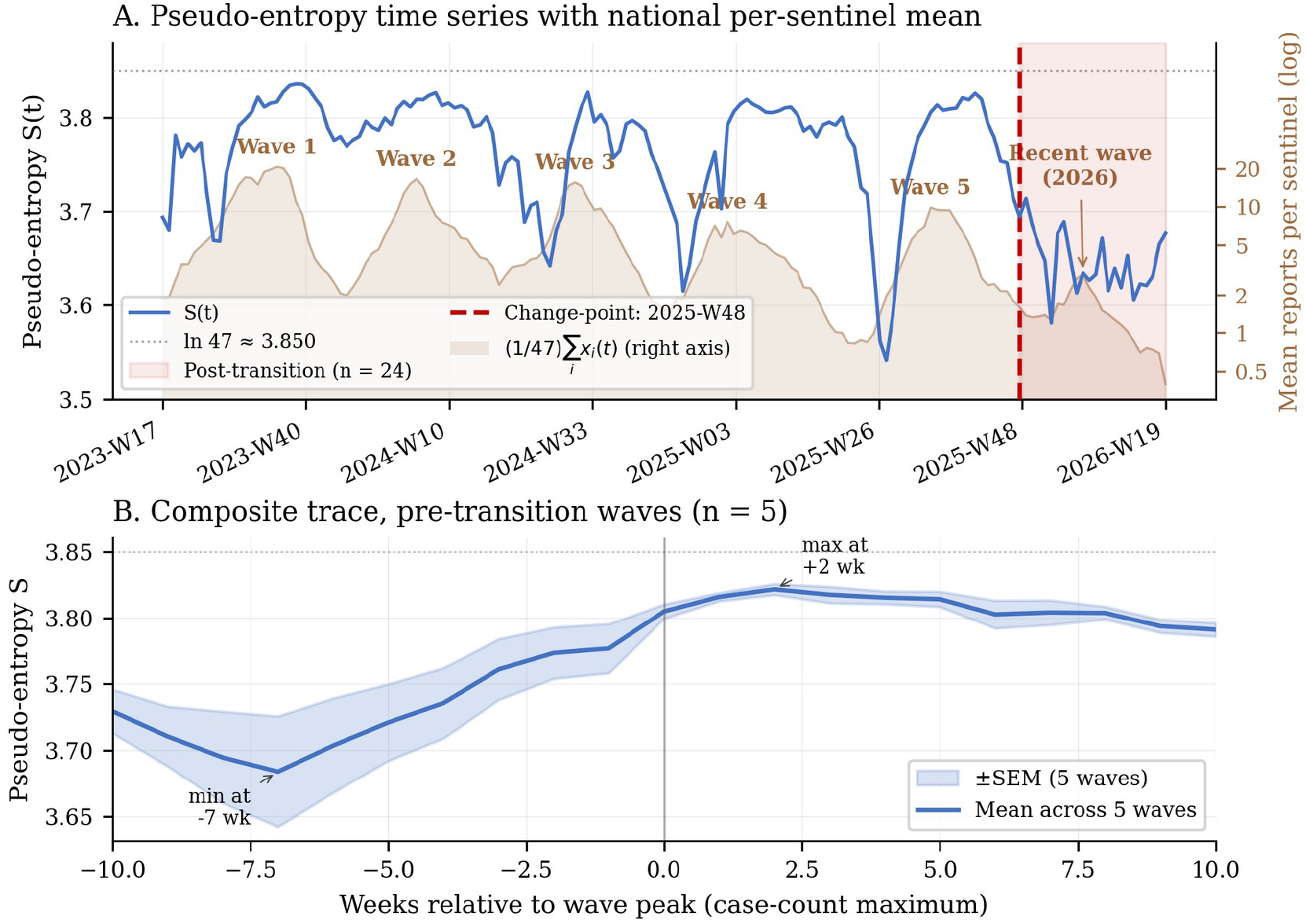
The post-Class-5 wave template. (A) Time series of the prefecture-share pseudo-entropy *S(t)* (blue, left axis) across all 159 weeks. The dotted reference line at ln 47 = 3.850 marks the maximum entropy. Local minima of *S* coincide with the inter-wave troughs of nationwide spatial concentration; local maxima occur during inter-wave plateaus. The unweighted national mean of weekly per-sentinel-facility reports, (1/47)Σ_i_x_i_(t), is overlaid as a shaded band on a logarithmic right-hand axis — the simple mean across the 47 prefectures (each weighted equally), distinct from the per-week normaliser Σ_j_x_j_(t) of Equation 1 — locating the five pre-transition waves (Waves 1–5) and the markedly smaller recent (2026) wave at their case-magnitude peaks. Wave amplitude declined monotonically across the pre-transition record (≈ 20.7 reports per sentinel at Wave 1 to ≈ 9.8 at Wave 5), while the recent wave reached only ≈ 2.9, falling within the post-transition period (red shading). The data-driven change-point at 2025-W48 is indicated by a vertical dashed line. (B) Composite trace of *S(t)* aligned to the case-count peak (t = 0) of the five pre-transition waves — the alignment reference now made directly visible by the per-sentinel curve in panel A — with the entropy minimum ∼7 weeks before and the entropy maximum ∼2 weeks after each peak.

### Wave-amplitude-invariant entropy ceiling

Although the nationwide peak amplitudes of the five pre-transition waves spanned a 2.7-fold range (from 7.6 to 20.7 reports per sentinel facility per week), the local maximum of *S(t)* attained during each post-peak inter-wave plateau was remarkably invariant: across the five waves *S*_max ranged only from 3.820 to 3.836 — a relative spread of 0.43% — and reached on average 99.4% of the theoretical upper bound ln 47 ≈ 3.850 (Table 1). The pre-transition *S*_max distribution was therefore tightly clustered with sample standard deviation 0.006, against which the post-transition maximum of 3.689 falls well outside the entire pre-transition range (see § “The post-transition departure”). Despite the 2.7-fold range in absolute wave amplitude, all five post-peak *S* maxima therefore fell within a narrow interval close to the theoretical upper bound, indicating that pseudo-entropy probes a feature of the post-peak spatial distribution that does not scale with wave amplitude.

This wave-amplitude-invariance of the *S* ceiling admits a natural empirical interpretation. The pseudo-entropy approaches its theoretical maximum ln 47 when prefecture-shares *p*_i_*(t) = x*_i_*(t)/*Σ_j_*x*_j_*(t)* approach uniformity. This can occur whenever the per-sentinel-facility case counts *x*_i_*(t)* undergo approximately uniform post-peak homogenisation across prefectures — whether through genuine saturation at a wave-specific level or through nationwide epidemic-wave propagation that equalises relative differences. Under either interpretation, the absolute level of *x*_i_ at homogenisation cancels in the ratio *p*_i_, so *S(t)* approaches ln 47 regardless of the wave’s absolute amplitude. We therefore refer to *S*_max as the *entropy ceiling* of the wave template, an empirical quantity defined by the inter-wave plateau of *S(t)*. The characteristic U-shape of the pre-transition *S* trajectory is asymmetric in its two limbs: the trough of *S* during wave growth is wave-amplitude-dependent (set by which prefecture is leading and by what factor), whereas the post-peak entropy ceiling is wave-amplitude-independent. This asymmetry is itself a testable feature of the wave template and motivates the interpretation of the post-transition ceiling drop in the next section.

### Data-driven identification of a structural break in late November 2025

Application of our two-stage change-point detection (Methods §“Change-point detection”) yielded a clear consensus (Figure 7). In Stage 1, the entropy ceiling at *S(t)* ≥ 3.80 was confirmed as a stable empirical regularity of the pre-transition record: all five pre-transition waves attained this threshold (Table 1, *S*_max range 3.820–3.836, mean 3.827, SD 0.006). In Stage 2, the last week in which *S(t)* attained the entropy ceiling was 2025-W42 (*S* = 3.820), corresponding to the post-peak plateau of the fifth pre-transition wave; *S(t)* did not return to the entropy ceiling in any subsequent week of the dataset.

Restricting the change-point search to the 29 subsequent weeks (2025-W43 to 2026-W19), PELT and CUSUM localised the break to late 2025 within a two-week window centred on late November 2025; Bai–Perron sup-F, with reduced power on the short restricted series, peaked later. PELT under any BIC-style penalty ≥ σ^2^·ln(*T*_r) identified 2025-W48 as the principal change-point; this estimate was invariant across the penalty range tested (0.5×–4× of the baseline). CUSUM produced its peak at 2025-W50 with empirical *p* < 0.0001 against the no-change null. Bai–Perron sup-F peaked at 2026-W02 with bootstrap *p* = 0.062, reflecting reduced statistical power on the short restricted series of *n* = 29 weeks. Sensitivity to the entropy-ceiling threshold choice across the range 3.78 ≤ threshold ≤ 3.82 — values that all fall below the pre-transition wave-ceiling minimum of 3.820 — yielded PELT change-points within the narrow range 2025-W47 – 2025-W49, that is, within ±1 week of the principal estimate of 2025-W48.

We therefore adopted 2025-W48 as the principal change-point and used it to define a *pre-transition* period (2023-W17 to 2025-W47; 135 weeks) and a *post-transition* period (2025-W48 to 2026-W19; 24 weeks) for all subsequent between-period analyses. Three features of this identification are noteworthy. First, the principal break is data-driven: no period was pre-specified as anomalous. Second, the identification is methodologically tight: the entropy ceiling is empirically established as a wave-amplitude-invariant feature of the pre-transition record, and the operational definition of the regime transition follows directly from this regularity rather than introducing a separate hypothesis. Third, the break is robust: PELT and CUSUM converge on a two-week window centred on late November 2025 (Bai–Perron sup-F, with reduced power, peaks later), and the PELT estimate is invariant across reasonable penalty values and across the natural range of entropy-ceiling thresholds.

A global change-point analysis applied to the full 159-week series (Supplementary Materials) reproduces 2025-W48 as the PELT estimate but additionally produces CUSUM and Bai-Perron primary peaks in early summer 2025 (2025-W22 and 2025-W25, respectively); inspection of the series shows that these earlier estimates are driven by a single-week transient dip in *S(t)* during the early growth phase of Wave 5, well before its August 2025 peak, rather than by the onset of a sustained shift. The restricted analysis described above naturally excludes these transient features.

### Two anti-phase spatial modes

Despite the apparent uniformity of the 5-wave template, Moran’s I revealed that low-*S* troughs corresponded to qualitatively distinct spatial topologies. Two prototypical low-*S* modes were identified (Figure 2): a *summer-onset, Okinawa-seeded Kyushu-cascade mode* (Mode A), in which low *S* coincides with low-to-moderate global Moran’s I, and a *winter-onset, Tohoku-centred connected-cluster mode* (Mode B), in which low *S* coincides with high global Moran’s I. The two modes differ qualitatively at the local-cluster level: Mode A is dominated by Okinawa as a geographically isolated high-low spatial outlier with onward seeding into a contiguous Kyushu-mainland cluster (Kagoshima, Miyazaki, Saga, Kumamoto, Nagasaki, Fukuoka, Oita — see Discussion and Supplementary Materials), while Mode B is dominated by a connected high-high cluster spanning Hokkaido, Aomori, Iwate, Miyagi, Akita, Yamagata, and Fukushima.

**Figure 2.**
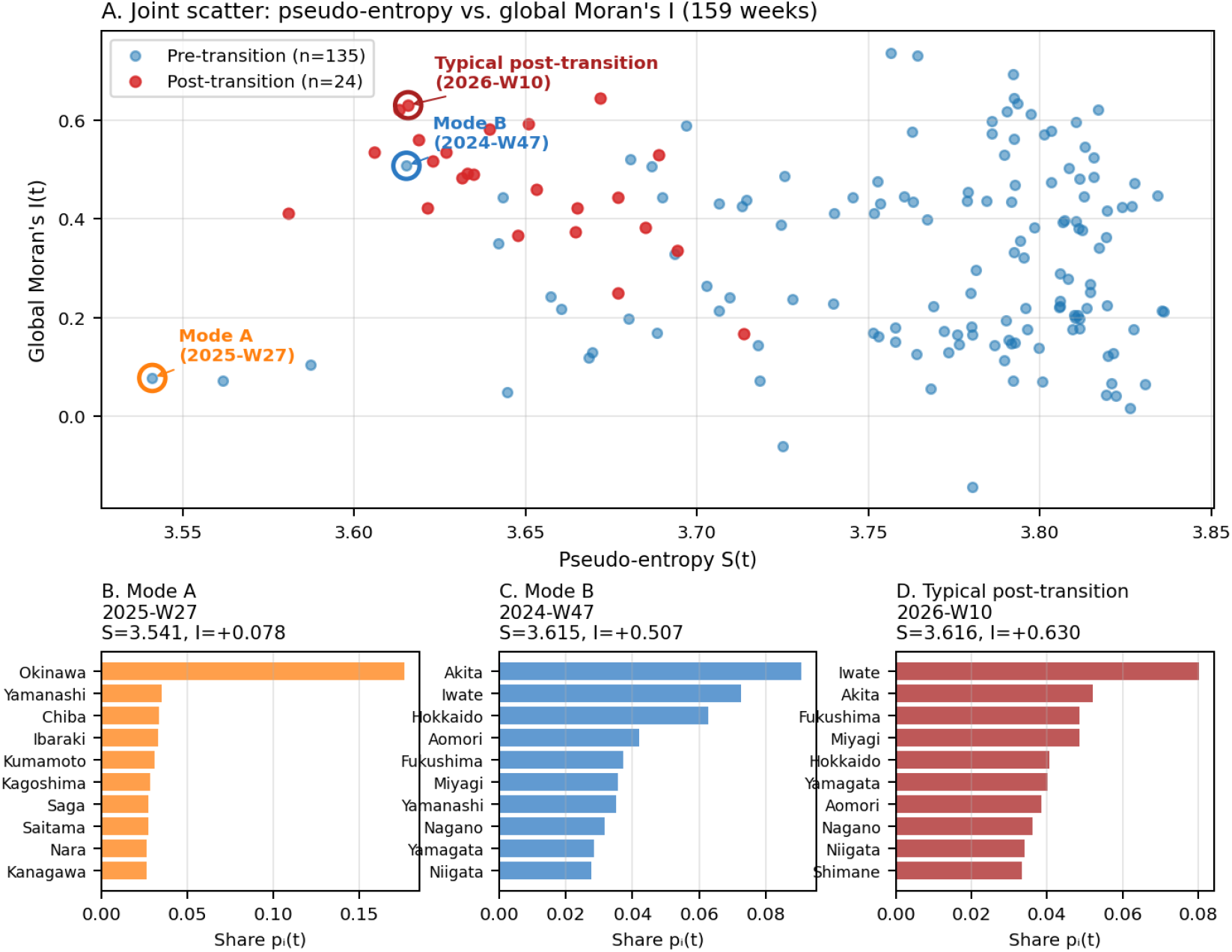
Identification of two qualitatively distinct spatial modes. (A) Joint scatter of pseudo-entropy *S(t)* and global Moran’s *I(t)* across all 159 weeks, colour-coded by pre-/post-transition. Three representative weeks are circled: Mode A (orange; 2025-W27), Mode B (blue; 2024-W47), and a typical post-transition week (dark red; 2026-W10). (B–D) Geographic visualization of the three representative weeks.

**Figure 3.**
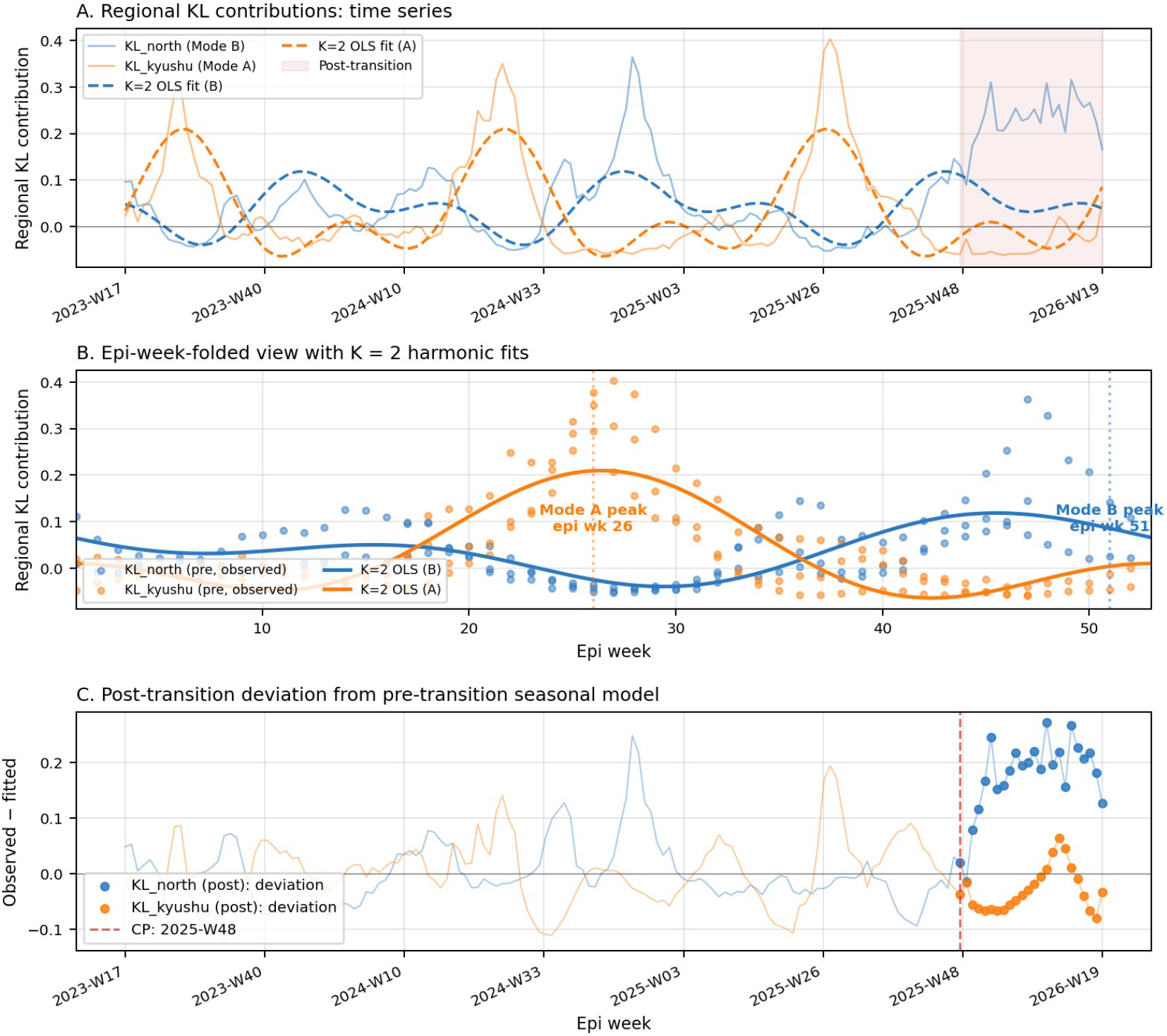
Seasonal anti-phase structure of Mode A and Mode B. (A) Time series of regional KL contributions: *KL_north* (Hokkaido + Tohoku, blue; Mode B signature) and *KL_kyushu* (Kyushu + Okinawa, orange; Mode A signature). Solid lines show observed values; dashed lines show second-order harmonic regression fits to the 135-week pre-transition data with AR(1) errors. The shaded red region marks the 24-week post-transition period. (B) Epi-week-folded view of the same data with overlaid harmonic fits. (C) Post-transition deviation from the seasonal model.

Second-order Fourier regression (equation 7, *K* = 2) of the regional KL contributions on the 135-week pre-transition data confirmed the seasonal-mode interpretation. Following common practice in environmental and epidemiological time-series analysis, we report the annual peak epi week from the OLS-fitted harmonic regression of the original-scale series, because the OLS coefficient estimates are unbiased estimators of the deterministic harmonic component and admit direct interpretation as the physical peak time of each mode. *KL_kyushu* (the principal signature of Mode A) had its annual peak at **epi week 26** (mid-Thursday 25 June); *KL_north* (the principal signature of Mode B) had its annual peak at **epi week 51** (mid-Thursday 16 December). The two modes were therefore separated by approximately 24.8 epi weeks in the annual cycle, within rounding of perfect anti-phase (26.1 epi weeks). The AR(1)-augmented (Cochrane–Orcutt) refit of the same model, with AR(1) coefficients 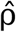 in the range 0.78–0.87 across these series (Supplementary Table S3), is used in §“Statistical analysis” to provide the residual covariance structure for inferential statistics; we do not use the AR(1)-innovation-space fit to assign the physical peak epi week, because the quasi-differencing transformation attenuates the slow deterministic annual component and is intended for inference rather than for descriptive characterisation of the seasonal cycle. The choice *K* = 2 was retained as the principal model because it captures the anti-phase decomposition with the minimum harmonic structure required by the data; sensitivity across *K* ∈ {1, …, 6} (Supplementary Materials) shows that the principal finding — the post-transition spatial-persistence regime transition described below — is invariant to *K*.

### The post-transition departure

The 24 post-transition weeks (2025-W48 to 2026-W19) exhibited three quantitative signatures of departure from the pre-transition template (Figure 4).

**Figure 4.**
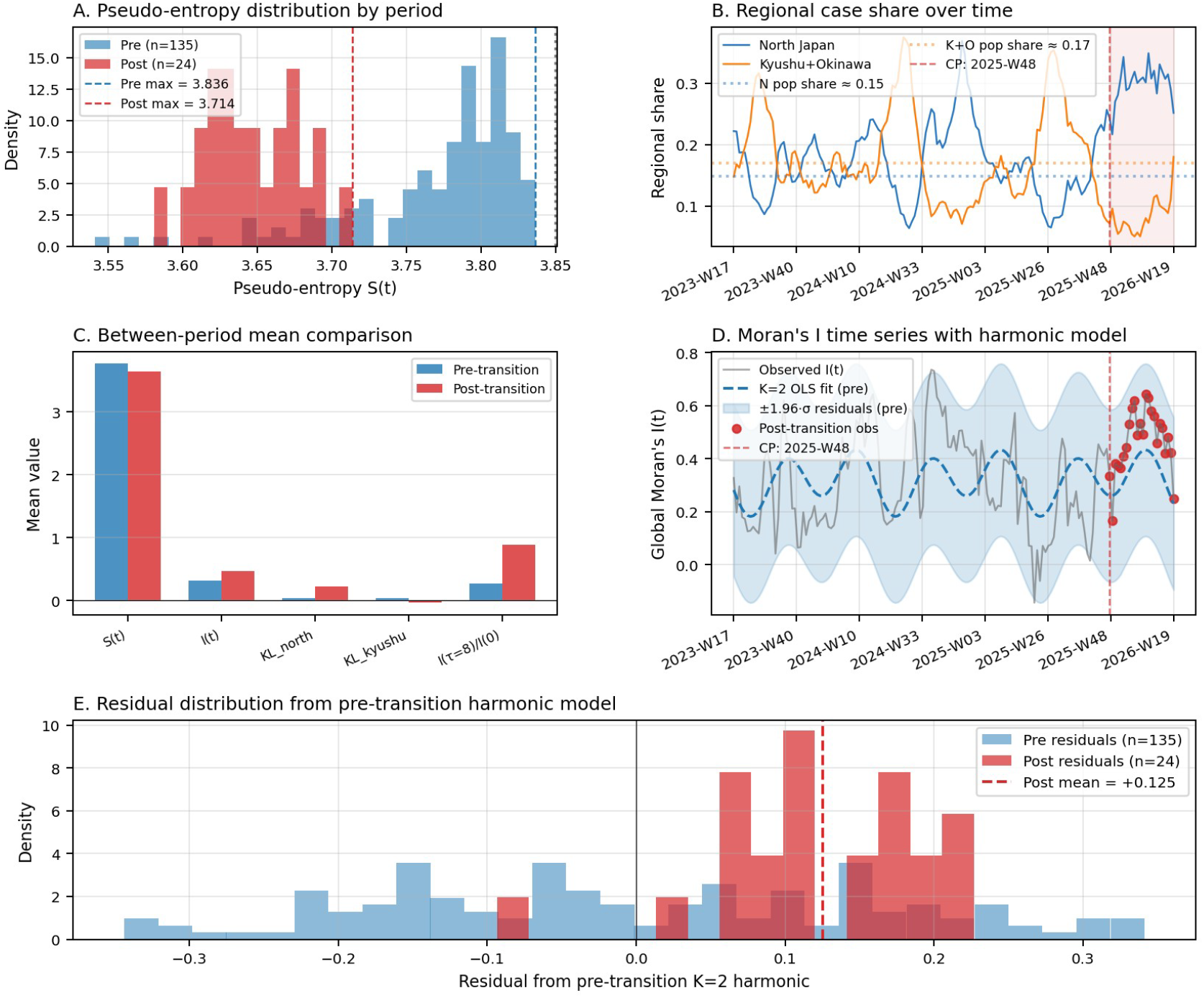
The post-transition departure. (A) Pseudo-entropy *S(t)* distribution: pre-transition (n=135) versus post-transition (n=24). The post-transition ceiling (max = 3.689) is well below the pre-transition ceiling (max = 3.836). (B) Aggregate share of cases attributable to North Japan (Hokkaido + Tohoku, blue) and Kyushu + Okinawa (orange), with population shares marked. (C) Between-period comparison of five quantitative indicators showing the magnitude of change. (D) Moran’s *I(t)* trajectory: grey line = observed, dashed blue = pre-transition second-order harmonic with AR(1) errors, shaded blue = 95% prediction band, red dots = post-transition observations. (E) Distribution of residuals from the pre-transition harmonic model.

#### Loss of the entropy ceiling

The maximum *S* attained during the post-transition period was 3.689 (2026-W18), corresponding to 95.8% of ln 47, well below the pre-transition entropy ceiling of 3.827 (SD 0.006). The mean *S* during the post-transition period was 3.658 (SD 0.027), compared with a pre-transition mean of 3.770 (SD 0.060), with a block-bootstrap mean difference of −0.112 (95% CI −0.142 to −0.082; *p* < 0.0001).

#### Regional redistribution

The mean share of cases attributable to Northern Japan (Hokkaido + Tohoku) rose from 0.158 in pre-transition to 0.295 in post-transition (Δ = 0.137; 95% bootstrap CI 0.092 to 0.181; *p* < 0.0001), while the share attributable to Kyushu + Okinawa fell from 0.187 to 0.097 (Δ = −0.090; 95% bootstrap CI −0.121 to −0.060; *p* < 0.0001). The pre-transition modal anti-phase between summer Kyushu+Okinawa dominance and winter Tohoku dominance was therefore not observed during the post-transition period; only the Tohoku/Northern signature was present.

#### Phase-plane signature

In the (*KL_north* − *KL_kyushu, I*) phase plane (Figure 5), pre-transition weeks traced an approximately closed orbit around the origin, consistent with seasonally alternating Modes A and B. All 24 post-transition weeks, in contrast, were confined to the upper-right quadrant (Mode B-dominated), with no excursion toward the lower-left (Mode A) quadrant.

**Figure 5.**
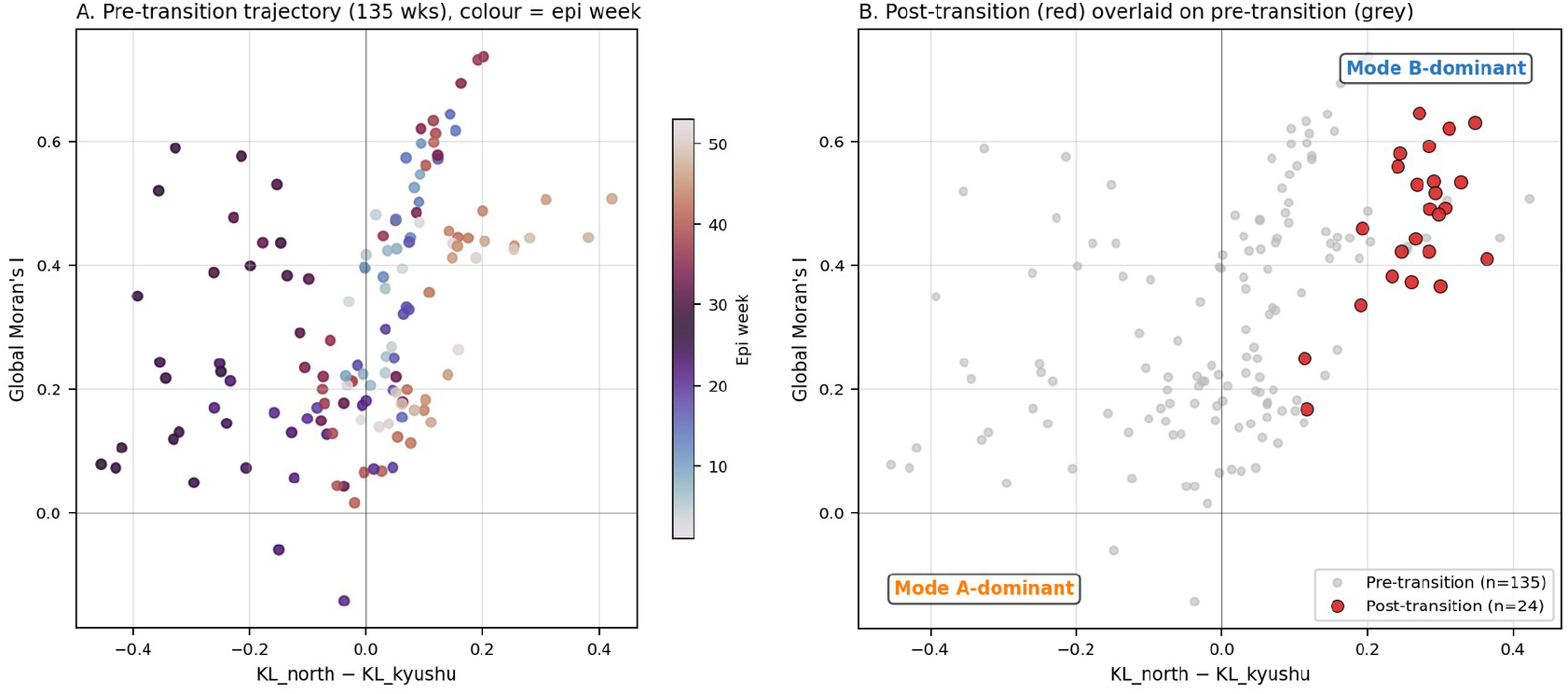
Phase-plane representation of the regime transition. The state is parameterized by the difference *KL_north* − *KL_kyushu* (horizontal axis) and global Moran’s *I* (vertical axis). (A) Pre-transition trajectory colour-coded by epi week (cyclic colourmap). (B) Post-transition trajectory (red filled circles) overlaid on pre-transition weeks (grey scatter). All 24 post-transition weeks are confined to the upper-right quadrant (Mode B-dominant), with no excursion toward the Mode A region.

### Spatial persistence of clusters: the principal signature of the regime transition

Time-lagged Moran’s I provides the most direct and methodologically robust evidence for the regime transition, because it measures temporal persistence of spatial structure independently of any time-series amplitude (Figure 6 and Table 2). Across all pre-transition weeks (n = 125 with positive *I*(0) at the corresponding lag), the spatial-persistence ratio *I(*τ *= 8)/I(0)* had a mean of 0.268 with a standard deviation of 0.840 — a highly variable distribution consistent with weak and inconsistent persistence of spatial clustering over 8 weeks, the canonical signature of traveling-wave epidemic dynamics. Over the 24 post-transition weeks, the corresponding ratio had a mean of 0.887 with a standard deviation of 0.338 — a tightly clustered distribution indicating that the same high-incidence prefectures and their geographic neighbours typically remained elevated 8 weeks later. The mean difference was 0.619 (95% bootstrap CI 0.315 to 0.897). Moving block bootstrap was significant across all block lengths tested: *b* = 1 (*z* = 6.15, *p* < 0.0001), *b* = 8 (*z* = 4.12, *p* < 0.0001), *b* = 12 (*z* = 4.08, *p* < 0.0001) (Supplementary Materials). The shift represents an approximately 3.3-fold elevation in mean spatial persistence at the 8-week lag, although we emphasise that, because the pre-transition mean is small relative to its standard deviation, the more interpretable summary is the mean shift of 0.62.

**Table 2.** Time-lagged Moran’s I and the spatial-persistence ratio, between-period comparison.

| Lag $\tau$ (wk) | Pre-transition, mean<br>(n) | Post-transition, mean<br>(n) | Persistence ratio<br>$I(\tau)/I(0)$ (pre; post) |
| --- | --- | --- | --- |
| 0 | 0.313 (135) | 0.468 (24) | 1.00; 1.00 |
| 1 | 0.300 (134) | 0.475 (24) | 0.96; 1.02 |
| 2 | 0.270 (133) | 0.472 (24) | 0.86; 1.01 |
| 4 | 0.186 (131) | 0.472 (24) | 0.59; 1.01 |
| 8 | 0.074 (127) | 0.418 (24) | 0.27; 0.89 |

The “post/pre” change in $I(\tau = 8)/I(0)$ : mean shift 0.62 (95% bootstrap CI 0.32 to 0.90); moving block bootstrap $p < 0.0001$ across block lengths 1–12. The persistence ratio for each lag is the mean of per-week ratios $I(\tau; t)/I(0; t)$ over weeks where $I(0; t) > 0$ .

**Figure 6.**
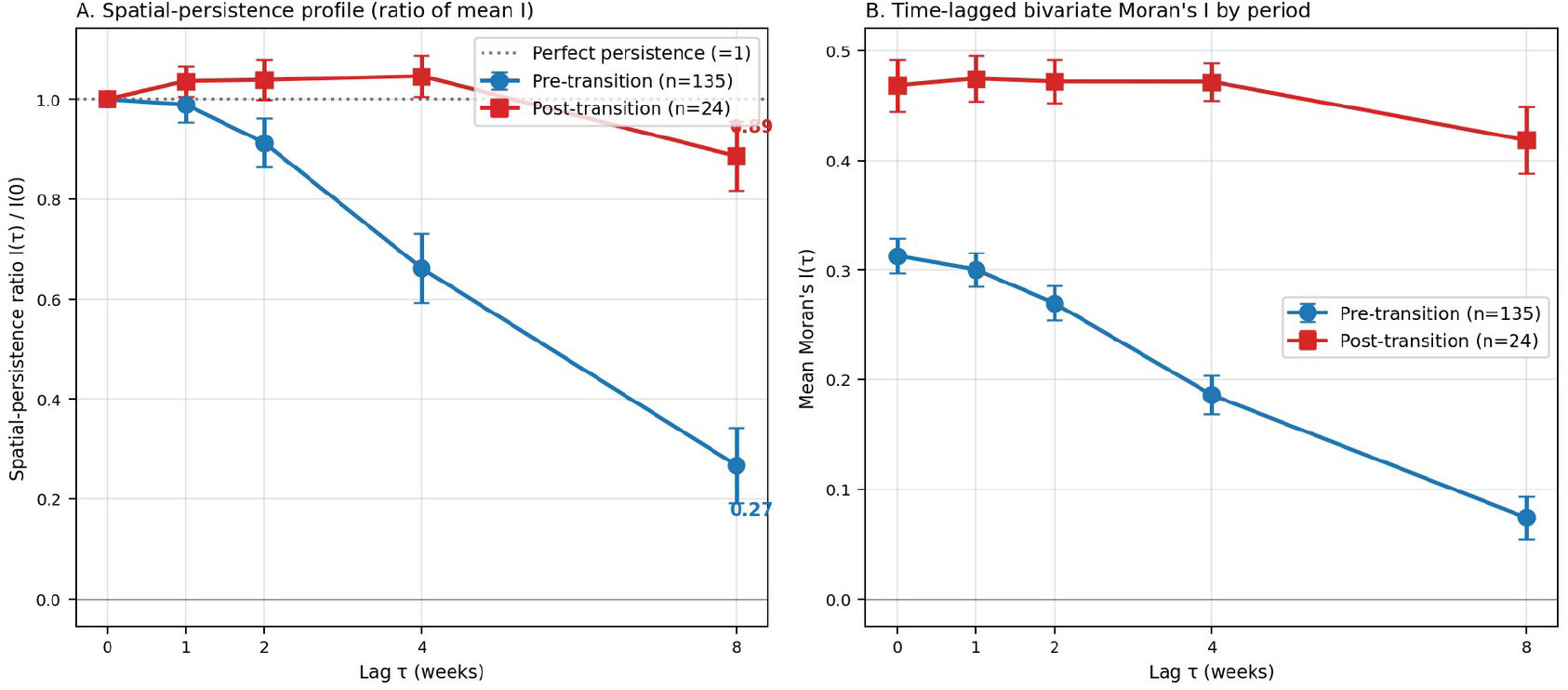
Time-lagged Moran’s I reveals persistent-clustering versus traveling-cluster regimes. (A) Spatial-persistence profile *I(*τ*)/I(0)* at lags τ ∈ {0, 1, 2, 4, 8} weeks. Pre-transition (n = 135, blue): the profile drops to ∼0.27 at τ = 8 weeks. Post-transition (n = 24, red): the profile remains near 0.89 at τ = 8 weeks. (B) Mean Moran’s *I* at each lag, period-stratified. Error bars are ±SE.

**Figure 7.**
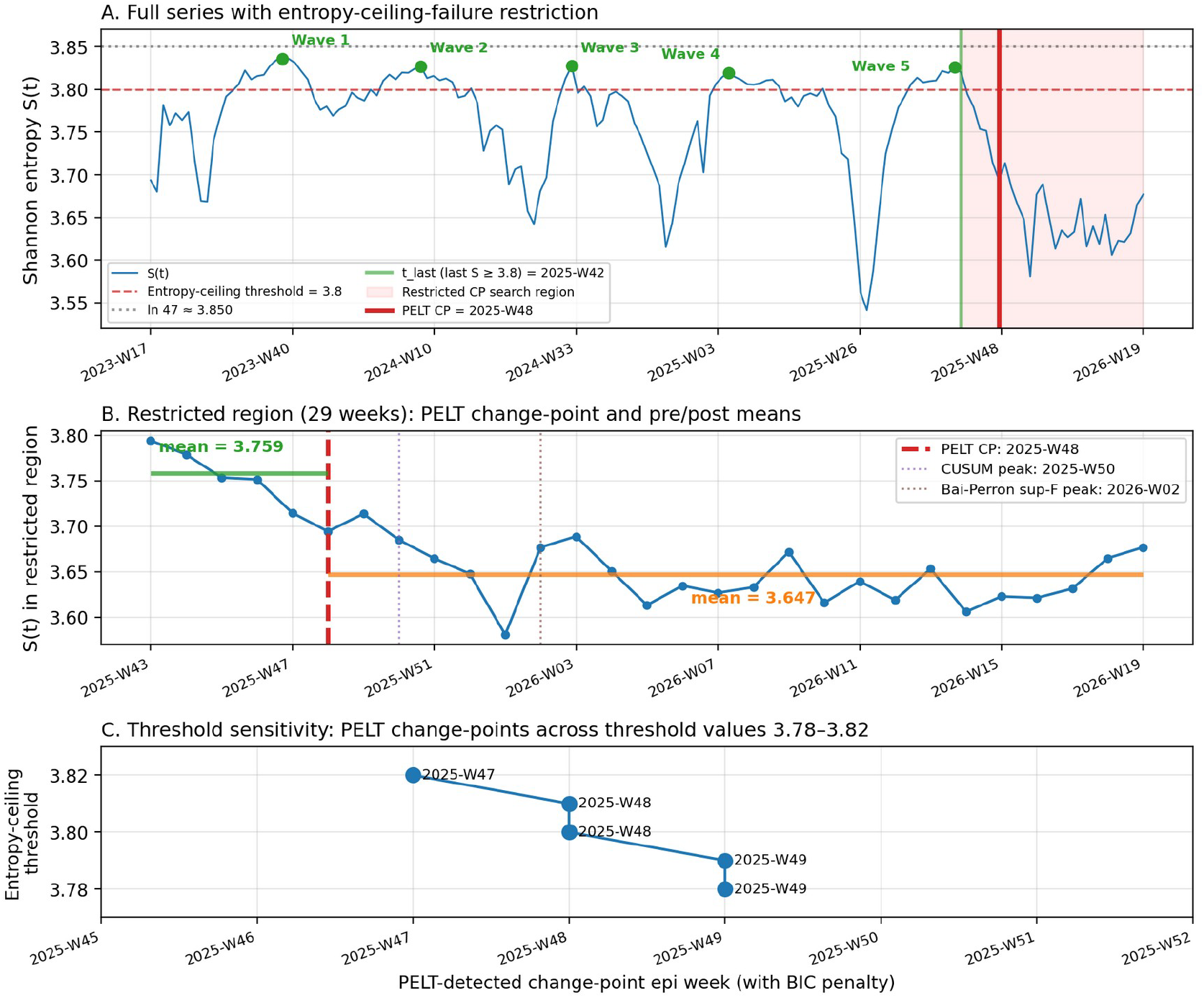
Data-driven entropy-ceiling-failure change-point detection. **(A)** Full 159-week time series of *S(t)* with the entropy-ceiling threshold of 3.80 (red dashed line) and the theoretical maximum ln 47 (grey dotted line). The five pre-transition wave maxima (Waves 1–5; green markers) all attained *S*_max ≥ 3.82, well above the entropy-ceiling threshold. The vertical green line marks the last week in which *S(t)* attained the entropy ceiling (t_last = 2025-W42, S = 3.820); the red-shaded region is the subsequent 29 weeks (2025-W43 to 2026-W19) used as the restricted change-point search region. The red vertical line marks the PELT-identified principal change-point at 2025-W48. **(B)** Restricted region zoomed: PELT change-point (red dashed line) cleanly separates a pre-CP mean of *S* = 3.759 (n = 5 weeks, green) from a post-CP mean of *S* = 3.647 (n = 24 weeks, orange). The CUSUM peak (2025-W50) and the Bai–Perron sup-F peak (2026-W02) are also shown as dotted vertical lines; all three methods agree within a two-week window centred on late November 2025. **(C)** Threshold sensitivity: PELT-detected change-points for entropy-ceiling thresholds 3.78 ≤ threshold ≤ 3.82 (all below the pre-transition wave-ceiling minimum of 3.820). Detected change-points fall within the narrow range 2025-W47 – 2025-W49, that is, within ±1 week of the principal estimate of 2025-W48.

Critically, this result holds under autoregressive-augmented null models. The spatial-persistence ratio *I*(τ)/*I(0)* is by construction the standardised temporal autocorrelation of the spatial pattern; an AR(1) null already accommodates persistence in the residuals of *I(t)*, but the observed shift from a pre-transition mean of 0.27 to a post-transition mean of 0.89 — a difference more than four bootstrap standard errors above zero — is extreme under any seasonal + AR(1) null in which the AR coefficient is estimated on pre-transition data. The spatial-persistence ratio therefore provides the most conservative and methodologically robust evidence for the regime transition.

### Sensitivity analyses

The principal findings — the late-November 2025 change-point and the post-transition elevation in spatial-persistence ratio — were robust to (i) alternative specifications of the spatial weights matrix **W**: across all seven spatial-weights specifications tested (including sparse network-based variants and a continuous distance-decay variant), the post-/pre-transition ratio of mean Moran’s I exceeded 1.35× in every variant (Supplementary Materials); (ii) harmonic-order choice *K* ∈ {1, …, 6}: AIC under AR(1)-augmented errors favours K ≥ 3 for the individual KL series (KL_kyushu, KL_north) but K = 2 captures the principal anti-phase decomposition relevant to the regime-transition claim, and the spatial-persistence ratio shift is invariant to *K* because it is computed directly from *I(*τ; *t)* without harmonic decomposition (Supplementary Materials); (iii) the April 2025 surveillance reform: restricting the principal between-period comparison to the post-reform period (n = 33 pre-transition weeks from epi week 15 of 2025 onwards, n = 24 post-transition weeks) yielded a mean shift of 0.58 with block-bootstrap *p* < 0.001, indicating that the regime transition is not an artefact of the reform; and (iv) Okinawa exclusion: re-computing all spatial-statistical quantities on the 46-prefecture subset preserved both the late-November 2025 change-point and the post-transition persistence elevation (Supplementary Materials).

## 4. Discussion

### Principal findings

In this 159-week ecological time-series analysis of prefecture-level COVID-19 sentinel surveillance data covering the entire post-Class-5 period in Japan, we document three principal findings. First, the five epidemic waves observed between August 2023 and August 2025 followed a stereotyped spatial trajectory in which the Shannon pseudo-entropy *S* of the prefecture-share distribution traced a characteristic U-shaped pattern around each wave peak a uniformity-then-concentration-then-recovery cycle that provides a quantitative template for the canonical post-Class-5 wave structure, characterised by a wave-amplitude-invariant entropy ceiling of *S*_max ≈ 99.4% of ln 47 (range 3.820–3.836, SD 0.006). Second, although superficially similar at the level of *S* alone, low-*S* troughs corresponded to qualitatively distinct spatial topologies, distinguishable only by the combined use of *S* and Moran’s I: a summer-onset Okinawa-seeded Kyushu cascade (Mode A; annual peak epi week 26) and a winter-onset Tohoku-centred connected cluster (Mode B; annual peak epi week 51), separated by approximately 25 epi weeks in the annual cycle — within rounding of perfect anti-phase.

Third, a two-stage data-driven change-point analysis — in which we used the pre-established entropy ceiling to define the operational onset of any regime transition as the last week in which *S(t)* attained this ceiling, and then restricted change-point search to subsequent weeks identified 2025-W48 as the principal structural break in the pseudo-entropy series. This estimate is robust across three independent change-point detection methods (PELT, CUSUM, Bai–Perron sup-F), across reasonable penalty specifications, and across the natural range of entropy-ceiling thresholds (3.78–3.82). The post-transition period exhibits three coherent signatures of departure from the pre-transition template — loss of the entropy ceiling, regional redistribution toward Northern Japan, and persistence of spatial clusters out to at least 8 weeks. The spatial-persistence ratio *I(*τ *= 8)/I(0)* shifted from a highly variable pre-transition distribution centred near 0.27 to a tightly clustered post-transition distribution around 0.89 (mean shift 0.62, 95% bootstrap CI 0.32 to 0.90; moving block bootstrap *p* < 0.0001), and this shift is the principal methodologically robust signature of the spatial-persistence regime transition. The methodological tightness of the change-point identification — that the operational definition follows directly from a pre-established empirical regularity (the entropy ceiling) rather than from a separately introduced hypothesis — strengthens the inferential basis for the regime-transition claim.

### Comparison with prior literature

Our findings extend three established lines of investigation in spatial epidemiology. The application of inequality and entropy indices to COVID-19 surveillance data was pioneered by Manz et al. [9] using country-level Theil and Gini indices, and was developed in our group’s earlier work along three complementary directions: an empirical phenomenological indicator (the K-value [6]); a mechanistic compartment model that explained why cumulative cases follow a Gompertz curve through the suppression of secondary and higher-order transmissions (the broken-link model [7]); and Theil-index decomposition of regional disproportionality applied to U.S. state-level data [8]. The pseudo-entropy *S* employed here is conceptually distinct from the Theil index — it measures deviation from spatial uniformity rather than from population-proportional distribution — but it occupies the same methodological niche as a single-number summary of spatial concentration. Combining *S* with Moran’s I, as we did here, addresses a known limitation of any global concentration index: that two regions with identical *S* or *T* can have profoundly different spatial topologies. The Mode A versus Mode B distinction is invisible to *S* alone but immediately apparent when *S* is paired with Moran’s I, mirroring the rationale for combining global with local indicators of spatial association advocated by Anselin [12].

Our two-mode finding is consistent with — and provides spatial-statistical support for — the prefecture-level reproduction-number heterogeneity reported by Wagatsuma et al. [5], who found that Okinawa exhibited the highest mean *R*_t_ across six selected prefectures during 2020– 2022, while Hokkaido exhibited the lowest. Our analysis extends this observation in two directions: it covers all 47 prefectures and the entire post-Class-5 period, and it characterizes the temporal behaviour of regional dominance rather than just static averages. Where Wagatsuma et al. observed asymmetry between Okinawa and Hokkaido, we show that this asymmetry is itself the signature of two seasonally distinct, anti-phased modes of geographic spread, with Okinawa seeding a Kyushu-mainland cascade in summer and the broader northern region anchoring winter outbreaks. The traveling-wave framework established for measles by Grenfell, Bjørnstad, and Kappey [15] and the metapopulation gravity-coupling framework of Xia, Bjørnstad, and Grenfell [17] provide the conceptual scaffolding for interpreting these modes as manifestations of distinct seasonal coupling structures.

Our third finding — the data-driven identification of a late-November 2025 regime transition with apparent absence of the Mode A signature during the post-transition period — connects to the broader question of pandemic-to-endemic transition. Theoretical work by Lavine, Bjornstad, and Antia [25] and Otto, MacPherson, and Colijn [26] predicts that as population immunity accumulates, SARS-CoV-2 dynamics should shift from epidemic to endemic with characteristic changes in age structure and seasonality. Our results suggest that this transition may also have a previously unrecognized spatial signature: the apparent suppression, or delayed re-emergence, of one of two seasonally distinct outbreak modes, leaving only the mode anchored to the structural drivers (in this case, winter respiratory pathology) most aligned with the broader endemic respiratory-virus calendar. The seasonality of respiratory and other infectious diseases is well documented and shaped by environmental and climatic drivers [27, 28, 29, 30].

### Mechanistic interpretation

A sensitivity analysis excluding Okinawa (Supplementary Materials) refined our initial characterisation of Mode A. In the 47-prefecture analysis, Mode A presented as a low-Moran’s-I summer pattern, suggesting geographic dispersion. However, when Okinawa was excluded, the Mode A summer-window Moran’s I rose substantially and exceeded that of Mode B, and inspection of prefecture-share rankings showed that the seven Kyushu-mainland prefectures (Kagoshima, Miyazaki, Saga, Kumamoto, Nagasaki, Fukuoka, Oita), each with a mean summer-window share above the national average, form a connected high-share cluster. We therefore reinterpret Mode A not as a point-source phenomenon but as an Okinawa-seeded Kyushu cascade: an isolated subtropical primary seed (Okinawa) followed by onward transmission into a contiguous Kyushu-mainland cluster. Mode B remains characterised as a Tohoku-centred connected cluster with no analogous seed-cascade structure. The regime transition identified in late November 2025 is robust to Okinawa exclusion, with both 47- and 46-prefecture analyses yielding qualitatively identical conclusions (post-transition spatial-persistence ratio elevated and Mode A signature not detected during the observed post-transition period).

### Implications for endemic transition theory

The observation that the post-transition spatial structure resembles a *spatially persistent cluster* — in which the same high-incidence prefectures and their geographic neighbours remain elevated over multi-week timescales — is qualitatively consistent with predictions from coupled-oscillator metapopulation theory [31, 32] in the strong-coupling or weak-forcing limit: when seasonal forcing weakens relative to inter-prefecture coupling, the system can transition from spatially synchronised epidemic waves [33] traversing the network (the *traveling-wave* regime) to spatially fixed clusters maintained by ongoing local transmission (the *persistent-clustering* regime). Such metapopulation approaches have been applied to forecast spatially structured epidemics in other settings [34]. Whether the late-November 2025 transition represents a genuine entry into this regime or a transient anomaly cannot be determined from the present 24-week observation window. We note specifically that the share of cases attributable to Okinawa, which had collapsed to 0.004 in early 2026 (2026-W02), has shown a steady increase over the most recent weeks of the dataset (0.046, 0.039, 0.034, 0.040, 0.045 in 2026-W15, 2026-W16, 2026-W17, 2026-W18, and 2026-W19, respectively), suggesting the possibility that Mode A is re-emerging rather than being structurally suppressed. Observation through summer 2026 is required to distinguish a sustained regime transition from a transient seasonal anomaly. Continued surveillance through summer 2026 will allow direct evaluation of whether the Mode A signature is structurally suppressed, delayed, or re-established under a persistent-clustering regime.

### Strengths and limitations

Strengths of this study include: (i) the use of all 47 prefectures and the entire 159-week post-Class-5 surveillance record, with no missing values; (ii) the principled combination of an information-theoretic concentration measure with spatial-autocorrelation statistics, enabling identification of seasonal modes invisible to either statistic alone; (iii) a fully data-driven approach to identifying the regime transition, with multi-method consistency providing robust evidence for a structural break in late November 2025; and (iv) the use of moving block bootstrap as the principal inferential statistic for all between-period comparisons, accommodating the strong serial dependence of weekly surveillance data.

Limitations include the following. First, the post-transition period is only 24 weeks at the time of analysis; whether the spatial-persistence regime transition is sustained or transient cannot be determined within this window, and longer follow-up is needed. Second, our analysis uses the prefecture as the unit of analysis, masking sub-prefectural heterogeneity (especially within larger prefectures such as Tokyo and Osaka); this is a fundamental limitation of the publicly available sentinel-surveillance data. Third, our methods are spatial-statistical and do not directly identify the mechanistic drivers of the regime transition — whether immunity accumulation, viral evolution, behavioural change, climatic factors, or surveillance-system effects (such as the April 2025 reform, which our sensitivity analysis suggests is not responsible) — and we explicitly avoid making causal claims about the mechanism. Fourth, the most recent weeks of the dataset (April–May 2026) show a partial re-emergence of Okinawa case shares, raising the possibility that the late-November 2025 transition is part of a longer cycle rather than a stable new regime; this is acknowledged above and is a major motivation for continued surveillance through summer 2026. Fifth, the change-point detection methods we use can identify *that* a structural break occurred and *when*, but do not identify the latent mechanism driving the break; mechanistic interpretation remains in the domain of mathematical-epidemiological modelling and is beyond the scope of the present study. Sixth, the dataset version used here, covering data through 2026-W19, was the most recent available at the time of analysis; updated analyses will become possible as further weekly reports accumulate.

## Conclusions

Across 159 weeks of post-Class-5 COVID-19 sentinel surveillance in Japan, the spatial structure of weekly case shares exhibits two anti-phased seasonal modes (a summer-onset Okinawa-seeded Kyushu cascade and a winter-onset Tohoku-centred connected cluster) and a wave-amplitude-invariant entropy ceiling. Data-driven change-point analysis identifies 2025-W48 as the principal structural break in the pseudo-entropy time series, and the subsequent 24-week post-transition period shows three coherent signatures of departure from the pre-transition template: loss of the entropy ceiling, regional redistribution, and elevated spatial persistence (with the 8-week persistence ratio increasing from 0.27 to 0.89). Whether this candidate spatial-persistence regime transition is sustained or transient remains to be determined; observation through summer 2026 is required, and the most recent uptick in Okinawa case shares leaves both interpretations open. The methodological framework developed here — combining information-theoretic, spatial-autocorrelation, and change-point statistics — provides a principled basis for ongoing surveillance of the spatial structure of endemic respiratory infections in Japan and, potentially, in other settings with comparable sentinel-surveillance systems.

## Supporting information

STROBE checklist

Supplementary adjacency edge list

## Statements

### Ethics statement

This study uses only publicly available aggregate prefecture-level surveillance data published by the Japan Ministry of Health, Labour and Welfare. No individual-level data were accessed. As an ecological analysis of fully aggregated, de-identified, public data, this study did not require institutional review board approval under Japanese ethical guidelines for medical research involving human subjects.

### Data availability

All input data are derived from publicly available MHLW reports at https://www.mhlw.go.jp/stf/seisakunitsuite/bunya/0000121431_00086.html. The processed prefecture-week dataset, the adjacency matrix, and the analysis code are openly available at https://github.com/takashi7nakano/covid-spatial-entropy-japan (archived at Zenodo, DOI: 10.5281/zenodo.20359405).

### Code availability

The complete analysis pipeline is implemented in Python 3.11 (NumPy 1.26, SciPy 1.12, pandas 2.2, scikit-learn 1.4, matplotlib 3.8, statsmodels 0.14, ruptures 1.1). Random permutation tests for Moran’s I use a fixed seed (np.random.seed(42)) for reproducibility. Code and processed data are openly available at https://github.com/takashi7nakano/covid-spatial-entropy-japan (archived at Zenodo, DOI: 10.5281/zenodo.20359405).

### Author contributions (CRediT)

T.N.: Conceptualization, methodology, data curation, formal analysis, writing — original draft, supervision, project administration. D.O.: Methodology, writing — review and editing. Y.I.: Methodology, formal analysis, writing — review and editing. K.W.: Methodology, writing — review and editing. Y.T.: Investigation, writing — review and editing. All authors approved the final manuscript.

## Acknowledgments

The authors thank the Japan Ministry of Health, Labour and Welfare and the National Institute of Infectious Diseases for maintaining the public surveillance data portal that made this analysis possible.

## Reporting guideline

This study is reported in accordance with STROBE (Strengthening the Reporting of Observational Studies in Epidemiology) guidelines for cross-sectional ecological studies. The completed STROBE checklist is provided as a separate supplementary file.

