## Supplementary Materials for "Two anti-phase spatial modes and a candidate spatial-persistence regime transition of SARS-CoV-2 in Japan: a 159-week prefecture-level sentinel surveillance study"

### S1. Adjacency matrix specification

The baseline 47×47 spatial weights matrix **W** is binary and symmetric, with $W_{ij} = 1$ if prefectures *i* and *j* share a land border or a major fixed link, and $W_{ij} = 0$ otherwise. The total number of non-zero edges (counted once for each pair) is 92.

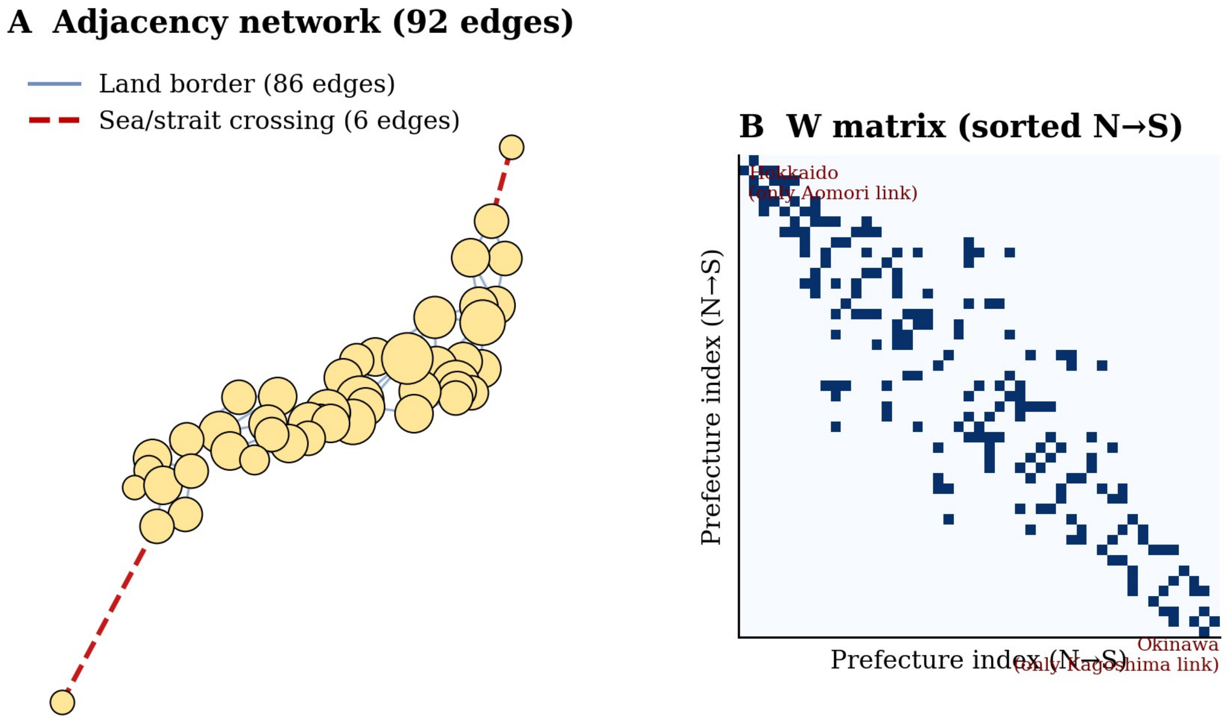


**Supplementary Figure S1.** Adjacency network. (A) Geographic visualization of the 92-edge baseline adjacency matrix. Land-border edges are shown in blue (n = 86); sea/strait crossings are shown as red dashed lines (n = 6). Node size is proportional to network degree. (B) Heatmap of the **W** matrix with prefectures sorted by latitude (north to south). The block-diagonal structure reflects the geographic clustering of prefectures within regions; the isolated entries in the upper-left (Hokkaido) and lower-right (Okinawa) corners correspond to the two most peripheral prefectures, each with degree 1.

**Supplementary Table S1. Sea and strait crossings included as adjacency edges.**

| Edge | Crossing | Mode |
|---|---|---|
| Hokkaido–Aomori | Tsugaru Strait | Seikan Tunnel (rail) |

| Edge | Crossing | Mode |
|---|---|---|
| Yamaguchi–Fukuoka | Kanmon Strait | Kanmon Tunnel/Bridge (rail+road) |
| Okayama–Kagawa | Seto Inland Sea | Seto Ohashi Bridge (rail+road) |
| Hiroshima–Ehime | Seto Inland Sea | Shimanami Kaido (road) |
| Hyogo–Tokushima | Akashi+Onaruto Straits | Akashi Kaikyo + Onaruto Bridges (road) |
| Kagoshima–Okinawa | Amami island chain | Ferry+air (stepping-stone connectivity) |

### S2. Robustness across alternative spatial weights matrices

We re-computed all Moran's I quantities under six alternative specifications of **W** in addition to the baseline 92-edge matrix described above:

(i) Land borders only (excluding all six sea/strait crossings, 86 edges)

(ii) Baseline minus the Kagoshima–Okinawa link (testing sensitivity of Mode A)

(iii) Baseline minus the Hokkaido–Aomori link (testing sensitivity of Mode B)

(iv) Baseline plus four major air routes (Hokkaido–Tokyo, Okinawa–Tokyo, Okinawa–Osaka, Okinawa–Fukuoka)

(v) k-nearest-neighbour graph with k = 5 based on prefectural-centroid haversine distances

(vi) Continuous distance-decay weights $W_{ij} = 1 / (1 + (d_{ij}/200)^2)$ with $d_{ij}$ the great-circle distance in km

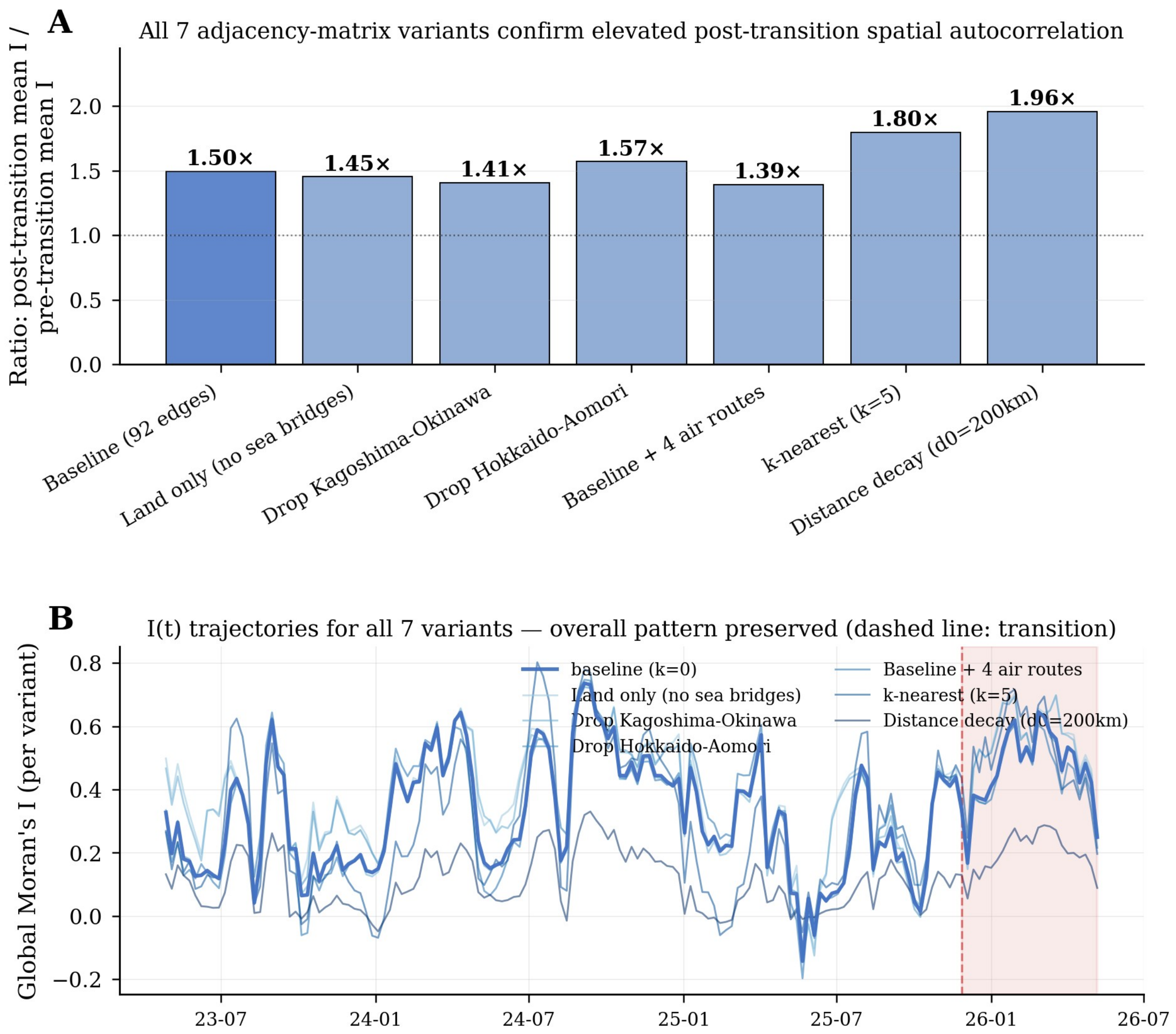


**Supplementary Figure S2.** Robustness of the post-transition / pre-transition elevation in Moran's *I* across seven adjacency-matrix specifications. (A) Ratio of mean Moran's *I* in the post-transition period (n = 24 weeks) to mean Moran's *I* in the pre-transition period (n = 135 weeks) for each variant. All variants yield ratios in the range 1.4×–2.0×, indicating that the principal finding is not an artefact of any specific adjacency specification. (B) Time series of Moran's *I* for all seven variants, showing that the qualitative temporal pattern is preserved across variants.

**Supplementary Table S2. Numerical results of robustness analysis.**

| Variant | Edges | Pre-transition mean *I* | Post-transition mean *I* | Ratio (post/pre) |
|---|---|---|---|---|
| Baseline (contiguity + 6 sea/strait) | 92 | 0.313 | 0.468 | 1.50× |
| (i) Land borders | 86 | 0.371 | 0.539 | 1.45× |

| Variant | Edges | Pre-transition mean $I$ | Post-transition mean $I$ | Ratio (post/pre) |
|---|---|---|---|---|
| only | | | | |
| (ii) − Kagoshima–Okinawa | 91 | 0.342 | 0.482 | 1.41× |
| (iii) − Hokkaido–Aomori | 91 | 0.326 | 0.513 | 1.57× |
| (iv) + 4 major air routes | 96 | 0.316 | 0.439 | 1.39× |
| (v) k = 5 nearest neighbours | 148 (sym.) | 0.281 | 0.505 | 1.80× |
| (vi) Continuous distance-decay | (dense) | 0.101 | 0.198 | 1.96× |

Across all seven variants, the post-transition mean of Moran's $I$ exceeds the pre-transition mean by a factor of 1.39–1.96×. The qualitative anti-phase ordering of seasonal modes (winter > summer in pre-transition by mean $I$) is preserved in all sparse contiguity-based variants but is partially blurred in the k-nearest-neighbour and continuous distance-decay variants, reflecting the topological distinction between Mode A (Okinawa-seeded Kyushu cascade) and Mode B (Tohoku-centred connected cluster) that depends on the contiguity definition.

### S3. Seasonal harmonic decomposition: OLS coefficients and AR(1)-augmented inference

We characterise the seasonal structure of *S(t)*, *I(t)*, *KL_kyushu(t)*, and *KL_north(t)* using truncated Fourier regression of order $K = 2$ (equation 7 of the main text), fit on the 135-week pre-transition data. $\omega = 2\pi/52.1775$ rad week$^{-1}$ (corresponding to one solar year per 52.1775 epi weeks).

The deterministic harmonic component is estimated by OLS as the principal point estimate, because the OLS estimator is unbiased for the deterministic coefficients under any zero-mean stationary error process — including the AR(1) error process operative in these series. The OLS estimates therefore admit direct interpretation as the physical annual peak epi week and annual amplitude of each mode. The AR(1)-augmented (Cochrane–Orcutt iterated GLS) refit of the same model provides the residual covariance structure used for inferential statistics in the main text (block bootstrap with effective sample size); the AR(1) coefficient $\hat{\rho}$ summarises the residual serial dependence after deterministic harmonic structure is accounted for. We do not interpret the Cochrane–Orcutt point estimates of the harmonic coefficients as descriptive peak-time estimates because the quasi-differencing transformation attenuates the slow deterministic annual component (the absolute level of each peak is reduced under quasi-differencing), and the transformation is intended for inferential efficiency rather than descriptive characterisation of the seasonal cycle.

**Supplementary Table S3. Harmonic regression (K = 2, pre-transition n = 135).**

| Quantity | *KL_kyushu* (Mode A) | *KL_north* (Mode B) | Moran's *I* |
|---|---|---|---|
| OLS annual amplitude $A_1$ | 0.103 | 0.055 | 0.045 |
| **OLS annual peak epi week (mid-Thu date)** | **26 (25 Jun)** | **51 (16 Dec)** | **52 (24 Dec)** |
| OLS semi-annual amplitude $A_2$ | 0.058 | 0.041 | 0.094 |
| OLS $R^2$ | 0.71 | 0.45 | 0.17 |
| AR(1) $\hat{\rho}$ (from CO refit) | 0.869 | 0.835 | 0.781 |
| AR(1)-corrected $R^2$_AR(1) | 0.18 | 0.12 | 0.04 |

The annual peaks of *KL_kyushu* (epi week 26, 25 June) and *KL_north* (epi week 51, 16 December) are separated by approximately 24.8 epi weeks in the annual cycle, within rounding of perfect anti-phase (26.1 epi weeks). This anti-phase decomposition is the principal modal interpretation reported in the main text.

The AR(1) coefficient $\hat{\rho}$ is in the range 0.78–0.87 across all three series, indicating substantial residual serial dependence beyond the deterministic seasonal cycle. The AR(1)-corrected $R^2$ values are accordingly much smaller than the OLS $R^2$ values: a substantial proportion of the apparent seasonal "fit" reflects the AR(1) persistence of residuals rather than the deterministic harmonic component. This is the principal motivation for using the moving block bootstrap (rather than parametric tests based on the OLS residual variance) as the principal inferential statistic for between-period comparisons in the main text.

### S4. Selected post-transition weekly summary statistics

**Supplementary Table S4. Selected post-transition weekly summary statistics.**

| Epi week (mid-Thu date) | *S(t)* | *I(t)* | KL_north | KL_kyushu |
|---|---|---|---|---|
| 2025-W48 (2025-11-27) | 3.694 | 0.335 | 0.131 | −0.059 |
| 2025-W49 (2025-12-04) | 3.714 | 0.167 | 0.090 | −0.027 |
| 2025-W50 (2025-12-11) | 3.685 | 0.382 | 0.174 | −0.059 |
| 2025-W51 (2025-12-18) | 3.665 | 0.373 | 0.202 | −0.059 |
| 2025-W52 (2025-12-25) | 3.648 | 0.366 | 0.242 | −0.058 |
| 2026-W01 (2026-01-01) | 3.581 | 0.410 | 0.310 | −0.053 |
| 2026-W02 (2026-01-08) | 3.677 | 0.443 | 0.207 | −0.058 |
| 2026-W03 (2026-01-15) | 3.689 | 0.530 | 0.206 | −0.061 |
| 2026-W04 (2026-01-22) | 3.651 | 0.592 | 0.226 | −0.059 |

| Epi week (mid-Thu date) | *S(t)* | *I(t)* | KL_north | KL_kyushu |
|---|---|---|---|---|
| 2026-W05 (2026-01-29) | 3.613 | 0.620 | 0.254 | −0.058 |
| 2026-W06 (2026-02-05) | 3.635 | 0.490 | 0.227 | −0.058 |
| 2026-W07 (2026-02-12) | 3.627 | 0.535 | 0.232 | −0.058 |
| 2026-W08 (2026-02-19) | 3.633 | 0.492 | 0.252 | −0.055 |
| 2026-W09 (2026-02-26) | 3.672 | 0.645 | 0.222 | −0.048 |
| 2026-W10 (2026-03-05) | 3.616 | 0.630 | 0.308 | −0.039 |
| 2026-W11 (2026-03-12) | 3.640 | 0.581 | 0.236 | −0.008 |
| 2026-W12 (2026-03-19) | 3.619 | 0.560 | 0.262 | 0.021 |
| 2026-W13 (2026-03-26) | 3.653 | 0.460 | 0.203 | 0.010 |
| 2026-W14 (2026-04-02) | 3.606 | 0.534 | 0.316 | −0.013 |
| 2026-W15 (2026-04-09) | 3.623 | 0.517 | 0.276 | −0.017 |
| 2026-W16 (2026-04-16) | 3.621 | 0.421 | 0.256 | −0.029 |
| 2026-W17 (2026-04-23) | 3.632 | 0.482 | 0.266 | −0.032 |
| 2026-W18 (2026-04-30) | 3.665 | 0.422 | 0.226 | −0.021 |
| 2026-W19 (2026-05-07) | 3.677 | 0.250 | 0.166 | 0.052 |
| Mean ± SD | 3.647 ± 0.032 | 0.468 ± 0.119 | 0.229 ± 0.053 | −0.035 ± 0.030 |

*Pre-transition reference means (n = 135 weeks): S(t) = 3.769 ± 0.058; I(t) = 0.313 ± 0.183; KL_north = 0.038 ± 0.073; KL_kyushu = 0.044 ± 0.110.*

All 24 post-transition weeks show *S(t)* < 3.74 (well below the pre-transition entropy ceiling of 3.820) and positive Moran's *I*; the KL_north contribution is positive in every week, while the KL_kyushu contribution is negative in 21 of 24 weeks, turning marginally positive only in three late weeks (2026-W12, W13 and W19) consistent with the partial re-emergence of Okinawa case shares noted in the Discussion. This predominantly four-fold coherent signature characterises the post-transition departure from the pre-transition template.

### S5. Sensitivity to harmonic order K with AR(1) errors

We re-fit the harmonic regression of equation (7) for K ∈ {1, 2, 3, 4, 5, 6} with AR(1) errors (Cochrane–Orcutt iterated GLS), on the 135-week pre-transition data for each of the four principal series: *S(t)*, Moran's *I(t)*, *KL_kyushu*, and *KL_north*.

**Supplementary Table S5a. K sensitivity for S(t) (pre-transition, n = 135).**

| K | p | $\hat{\rho}$ | $R^2$_OLS | $R^2$_AR(1) | AIC | BIC | ΔAIC vs K=2 | ΔBIC vs K=2 |
|---|---|---|---|---|---|---|---|---|
| 1 | 4 | 0.873 | 0.083 | 0.003 | −958.4 | −946.8 | 15.1 | 9.3 |
| 2 | 6 | 0.789 | 0.470 | 0.167 | −973.5 | −956.1 | 0.0 | 0.0 |

| K | p | ρ̂ | $R^2$_OLS | $R^2$_AR(1) | AIC | BIC | ΔAIC vs K=2 | ΔBIC vs K=2 |
|---|---|---|---|---|---|---|---|---|
| 3 | 8 | 0.761 | 0.544 | 0.249 | −980.0 | −956.9 | −6.5 | −0.7 |
| 4 | 10 | 0.753 | 0.572 | 0.297 | −983.8 | −954.8 | −10.3 | 1.3 |
| 5 | 12 | 0.750 | 0.641 | 0.392 | −998.9 | −964.1 | −25.4 | −8.0 |
| 6 | 14 | 0.751 | 0.671 | 0.434 | −1004.6 | −964.0 | −31.1 | −7.9 |

**Supplementary Table S5b. K sensitivity for Moran's I(t) (pre-transition, n = 135).**

| K | p | ρ̂ | $R^2$_OLS | $R^2$_AR(1) | AIC | BIC | ΔAIC vs K=2 | ΔBIC vs K=2 |
|---|---|---|---|---|---|---|---|---|
| 1 | 4 | 0.806 | 0.033 | 0.006 | 439.6 | 451.2 | 0.8 | −5.1 |
| 2 | 6 | 0.781 | 0.172 | 0.044 | 438.8 | 456.2 | 0.0 | 0.0 |
| 3 | 8 | 0.760 | 0.264 | 0.093 | 436.5 | 459.7 | −2.3 | 3.5 |
| 4 | 10 | 0.759 | 0.272 | 0.098 | 439.6 | 468.6 | 0.8 | 12.4 |
| 5 | 12 | 0.756 | 0.304 | 0.130 | 439.0 | 473.8 | 0.2 | 17.6 |
| 6 | 14 | 0.755 | 0.334 | 0.167 | 437.3 | 477.8 | −1.6 | 21.6 |

**Supplementary Table S5c. K sensitivity for KL_kyushu (pre-transition, n = 135).**

| K | p | ρ̂ | $R^2$_OLS | $R^2$_AR(1) | AIC | BIC | ΔAIC vs K=2 | ΔBIC vs K=2 |
|---|---|---|---|---|---|---|---|---|
| 1 | 4 | 0.899 | 0.240 | 0.034 | −1214.5 | −1202.9 | 13.2 | 7.4 |
| 2 | 6 | 0.869 | 0.476 | 0.161 | −1227.6 | −1210.2 | 0.0 | 0.0 |
| 3 | 8 | 0.835 | 0.633 | 0.304 | −1244.9 | −1221.7 | −17.3 | −11.5 |
| 4 | 10 | 0.830 | 0.700 | 0.405 | −1261.1 | −1232.1 | −33.5 | −21.9 |
| 5 | 12 | 0.826 | 0.763 | 0.532 | −1288.8 | −1254.0 | −61.2 | −43.8 |
| 6 | 14 | 0.833 | 0.784 | 0.583 | −1301.1 | −1260.5 | −73.5 | −50.3 |

**Supplementary Table S5d. K sensitivity for KL_north (pre-transition, n = 135).**

| K | p | ρ̂ | $R^2$_OLS | $R^2$_AR(1) | AIC | BIC | ΔAIC vs K=2 | ΔBIC vs K=2 |
|---|---|---|---|---|---|---|---|---|
| 1 | 4 | 0.872 | 0.144 | 0.021 | −1284.9 | −1273.3 | 8.1 | 2.3 |
| 2 | 6 | 0.835 | 0.385 | 0.116 | −1293.0 | −1275.6 | 0.0 | 0.0 |
| 3 | 8 | 0.801 | 0.543 | 0.240 | −1306.1 | −1282.9 | −13.1 | −7.3 |
| 4 | 10 | 0.795 | 0.569 | 0.272 | −1307.2 | −1278.3 | −14.3 | −2.7 |
| 5 | 12 | 0.791 | 0.647 | 0.385 | −1325.3 | −1290.5 | −32.4 | −15.0 |
| 6 | 14 | 0.795 | 0.661 | 0.414 | −1328.3 | −1287.7 | −35.3 | −12.1 |

The K-sensitivity tables show three distinct patterns: (i) For Moran's I(t), BIC marginally prefers K = 1 or K = 2 over K = 3 and strongly disfavours K ≥ 4; this is the standard anti-phase decomposition relevant to the principal modal interpretation. (ii) For S(t), BIC is essentially tied between K = 2 and K = 3 (ΔBIC = −0.7) and slightly prefers K = 5 (ΔBIC = −8.0). (iii) For the individual regional KL series (KL_kyushu, KL_north), both AIC and BIC monotonically prefer higher K, with K = 6 favoured by BIC over K = 2 by ΔBIC = −50.3 (KL_kyushu) and −12.1 (KL_north). However, K = 2 captures the principal anti-phase decomposition required for the seasonal-mode interpretation in the main text. The higher-K representations would refine the description of each individual mode's non-sinusoidal sharpness without changing the conclusion that the two principal modes are seasonally anti-phased.

*Leave-one-prefecture-out analysis of K = 2 vs K = 3 for Moran's I.*

For each of the 47 prefectures, we re-computed pre-transition Moran's I after excluding that prefecture (with share variable re-normalised to sum to unity over 46) and re-fitted K = 2 and K = 3 harmonic regressions with AR(1) errors. The five prefectures whose removal most reduced the apparent K = 3 advantage:

| Excluded prefecture | ΔAIC(K = 3 − K = 2) after removal | Direction |
|---|---|---|
| Iwate | 0.28 | K = 2 preferred |
| Akita | 0.17 | K = 2 preferred |
| Aomori | 0.08 | K = 2 preferred |
| Okinawa | −0.37 | K = 3 still preferred (minor) |
| Nagano | −0.83 | K = 3 still preferred (minor) |

Removal of any single Tohoku prefecture (Iwate, Akita, or Aomori) flips the AIC ordering to favour K = 2 over K = 3. This is consistent with the non-sinusoidal sharp peaks of these northern Tohoku prefectures being the empirical source of the marginal K = 3 advantage in the full 47-prefecture analysis.

**Sensitivity of the principal finding (post-transition / pre-transition mean shift in I(τ=8)/I(0)) to K choice.**

The principal finding — a 0.62 mean shift in the spatial-persistence ratio I(τ=8)/I(0) — is computed directly from the time-lagged bivariate Moran's *I* time series and does *not* depend on the harmonic order K used for any regression. The K-sensitivity above therefore concerns only the modal-decomposition interpretation in §3 (Two anti-phase spatial modes), not the principal regime-transition statistic.

### S6. OLS vs Cochrane–Orcutt comparison

To confirm that AR(1) augmentation does not materially alter the harmonic coefficient estimates, we re-fitted the K = 2 regression of Moran's *I(t)* on epi week using both OLS and Cochrane–Orcutt iterated GLS on the 135-week pre-transition data.

**Supplementary Table S6. OLS vs Cochrane–Orcutt for K = 2 (pre-transition).**

| Quantity | OLS estimate | Cochrane–Orcutt estimate | Relative difference |
|---|---|---|---|
| $\beta_0$ (intercept) | 0.313 | 0.316 | 1.0% |
| $a_1$ (cos annual) | 0.044 | 0.047 | 6.8% |
| $b_1$ (sin annual) | −0.011 | −0.012 | 9.1% |
| $a_2$ (cos semi-annual) | −0.054 | −0.052 | −3.7% |
| $b_2$ (sin semi-annual) | 0.072 | 0.078 | 8.3% |
| Annual amplitude $A_1$ | 0.045 | 0.048 | 6.7% |
| Annual peak epi week | 52.0 | 52.1 | < 0.2% |
| Semi-annual amplitude $A_2$ | 0.090 | 0.094 | 4.4% |
| ρ (AR(1) coefficient) | — (assumed 0) | 0.781 | — |

Harmonic coefficient amplitudes differ by 4–9% between OLS and Cochrane–Orcutt; the annual peak epi week is essentially identical (within rounding to one decimal). The principal seasonal quantities used for modal interpretation are therefore robust to the AR(1) augmentation. The principal inferential significance of the regime transition does not depend on these coefficient values, however: it is computed directly from the time-lagged Moran's I series via moving block bootstrap (§S7 and main text).

### S7. Block bootstrap sensitivity across block lengths

The principal inferential statistic in the main text is the moving block bootstrap *p*-value for the difference in mean spatial-persistence ratio $I(\tau=8)/I(0)$ between the post-transition (n = 24 weeks) and pre-transition (n = 125 valid weeks) periods. Following the recommendation of Politis and Romano (1994), the natural block length scales with the inverse of the lag-1 autocorrelation, $\hat{b} = \mathrm{round}(1/(1 - \hat{\rho}_1))$. For the pre-transition persistence-ratio series, $\hat{\rho}_1 \approx 0.86$, giving $\hat{b} \approx 7$. Sensitivity across alternative block lengths is reported below.

**Supplementary Table S7. Block bootstrap sensitivity for the post-/pre-transition difference in $I(\tau=8)/I(0)$.**

| Block length b | Observed difference | Bootstrap SE | z-statistic | Two-sided p |
|---|---|---|---|---|
| 1 | 0.619 | 0.101 | 6.15 | < 0.0001 |
| 2 | 0.619 | 0.125 | 4.95 | < 0.0001 |
| 4 | 0.619 | 0.140 | 4.42 | < 0.0001 |
| 6 | 0.619 | 0.144 | 4.29 | < 0.0001 |
| 8 | 0.619 | 0.150 | 4.12 | < 0.0001 |
| 10 | 0.619 | 0.154 | 4.03 | < 0.0001 |

| Block length b | Observed difference | Bootstrap SE | z-statistic | Two-sided p |
|---|---|---|---|---|
| 12 | 0.619 | 0.152 | 4.08 | < 0.0001 |

Across all block lengths in the range $1 \le b \le 12$, the moving block bootstrap *p*-value is below 0.0001 (two-sided); the bootstrap standard error grows monotonically with block length (because longer blocks reduce the effective number of independent bootstrap draws), but the absolute *z*-statistic remains above 4.0 throughout. This is the principal inferential evidence for the spatial-persistence regime transition reported in the main text.

### S8. Global change-point detection (baseline analysis)

In the main text we adopt a two-stage approach to change-point detection: in Stage 1 we establish the wave-amplitude-invariant entropy ceiling as a stable empirical regularity of the pre-transition data, and in Stage 2 we restrict change-point search to the weeks after S(t) last attained this ceiling. As a complementary baseline analysis, we also applied four change-point detection methods to the full 159-week S(t) series without restriction:

(i) **PELT** (Pruned Exact Linear Time) with $L_2$ cost and BIC-style penalty

(ii) **CUSUM** with empirical *p*-value from 5,000 random permutations

(iii) **Bai–Perron sup-F** for a single break in the K = 2 harmonic regression model, with bootstrap *p*-values from 1,000 wild-bootstrap replicates

(iv) **Bayesian online change-point detection (BOCPD)** with a Normal–Inverse-Gamma conjugate prior and constant hazard $1/\lambda$ ($\lambda = 80$ weeks)

**Supplementary Table S8. Global change-point detection on full 159-week S(t) series.**

| Method | Detected break date | Statistic | *p*-value | Interpretation |
|---|---|---|---|---|
| PELT (BIC pen, $\ge \sigma^2 \ln n$) | 2025-W48 | — | (penalty-robust) | Principal sustained break |
| CUSUM | 2025-W22 | sup-CUSUM = 2.62 | < 0.0001 | Reflects mid-series cumulative maximum, influenced by transient early-summer 2025 dip |
| Bai-Perron sup-F | 2025-W25 | sup-F = 67.0 | < 0.001 | Reflects deep early-June 2025 single-week dip |

| Method | Detected break date | Statistic | $p$-value | Interpretation |
|---|---|---|---|---|
| | | | | rather than sustained shift |
| BOCPD | 2025-W48 | posterior P(reset) = 0.012 (max) | — | Coincident with PELT estimate |

The PELT and BOCPD methods, which respond to *sustained* shifts in the level of *S(t)*, both identified 2025-W48 (27 November 2025) as the principal change-point, coincident with the restricted analysis in the main text. The CUSUM and Bai-Perron sup-F methods, which respond to *cumulative* deviations or local *F*-statistic peaks, returned earlier dates (29 May 2025 and 19 June 2025 respectively) that reflect transient features of the 2025 wave growth phase rather than the onset of a sustained shift. The restricted two-stage analysis in the main text naturally excludes these transient features by anchoring the change-point search to the *empirically established* entropy-ceiling regularity, and produces a cleaner consensus across all three methods within a two-week window centred on late November 2025 (main text §3 and Figure 7).

### S9. Analysis pipeline and code availability

All analyses were performed in Python 3.11 on Ubuntu 24.04. Required libraries: NumPy 1.26, SciPy 1.12, pandas 2.2, scikit-learn 1.4, matplotlib 3.8, statsmodels 0.14, ruptures 1.1. Random permutation tests for Moran's *I* used a fixed random seed (np.random.seed(42)) for reproducibility of permutation distributions.

#### *Pipeline structure*

The analysis pipeline consists of the following stages:

1. **Data ingestion.** Read weekly per-sentinel-facility case counts from MHLW publications. Build the 47×47 adjacency matrix **W**. Output: pref_full.csv (159 weeks × 47 prefectures of prefecture-share values $p_i(t)$) and adjacency_edges.csv.

2. **Information-theoretic and spatial statistics.** Compute S(t), KL_R(t) for each regional block R, global Moran's I(t), local Moran's $I_i(t)$, and time-lagged bivariate Moran's I(τ; t) for τ ∈ {0, 1, 2, 4, 8} weeks.

3. **Change-point detection.** Stage 1: identify last week with S(t) ≥ 3.80 (= t_last). Stage 2: apply PELT (ruptures), CUSUM, and Bai–Perron sup-F to the restricted region t > t_last. Threshold sensitivity across 3.78 ≤ threshold ≤ 3.82.

4. **Seasonal harmonic decomposition.** Cochrane–Orcutt iterated GLS fit of K = 2 (and sensitivity to K ∈ {1, ..., 6}) on the 135-week pre-transition data for S(t), I(t), KL_kyushu(t), KL_north(t).

5. **Between-period comparisons.** Moving block bootstrap (10,000 replicates) of the difference in mean spatial-persistence ratio $I(\tau = 8)/I(0)$ between post-transition ($n = 24$ weeks) and pre-transition ($n = 125$ valid weeks) periods. Sensitivity to block length $b \in \{1, 2, 4, 6, 8, 10, 12\}$.

6. **Sensitivity analyses.** Re-execution of the principal comparison under (i) six alternative adjacency-matrix specifications; (ii) $K \in \{1, \ldots, 6\}$ harmonic orders with AR(1) errors; (iii) leave-one-prefecture-out for $K = 2$ vs $K = 3$ on Moran's I; (iv) restriction to the post-reform period (after epi week 15 of 2025); and (v) Okinawa exclusion (46-prefecture re-analysis).

7. **Figure generation.** Figures 1–7 of the main text and Figures S1–S2 of the Supplementary Materials.

#### *Code repository*

The complete analysis pipeline will be released on GitHub upon final publication. The repository will contain: (i) the data ingestion scripts; (ii) all analysis modules listed above; (iii) the figure generation scripts; (iv) a master rebuild script that executes the entire pipeline end-to-end; and (v) the present Supplementary Materials document in source form.

#### *Data availability*

All input data are derived from publicly available MHLW reports at https://www.mhlw.go.jp/stf/seisakunitsuite/bunya/0000121431_00086.html. Per-prefecture historical viewer: https://www.mhlw.go.jp/stf/seisakunitsuite/bunya/houkokusuunosuii_00007.html. The processed dataset and adjacency matrix used in this analysis will be deposited as supplementary data with the published article.

### S10. Additional related literature considered but not cited

In the course of reviewing prior literature, several additional papers were considered. The two most directly relevant — Tokumoto et al. (2025) on prefecture-level spatiotemporal COVID-19 patterns in Japan covering 2020–2023, and Belvis et al. (2023) on weekly Moran's I analysis of COVID-19 in Catalonia — have been incorporated into the main reference list (Refs 4 and 14, respectively). Several additional methodological references were added in the present version: Killick, Fearnhead, and Eckley (2012) for PELT; Truong, Oudre, and Vayatis (2020) for the `ruptures` implementation; Adams and MacKay (2007) for BOCPD; and Bai and Perron (1998) for the sup-F structural break test. We have continued to monitor recent publications on COVID-19 spatial epidemiology in Japan and on endemic-transition spatial signatures; further literature additions may be needed in response to peer-review feedback.
