## Supplementary material for "Two anti-phase spatial modes and a candidate spatial-persistence regime transition of SARS-CoV-2 in Japan: a 159-week prefecture-level sentinel surveillance study": STROBE checklist

**STROBE Statement — Checklist for cross-sectional ecological studies**

*Reference: Vandenbroucke JP et al. (2007). STROBE: explanation and elaboration. Ann Intern Med 147(8): W163–94.*

| Section / Topic | Item # | Recommendation | Page / Section in our manuscript |
|---|---|---|---|
| **Title and abstract** | | | |
| | 1 | (a) Indicate the study's design with a commonly used term in the title or the abstract | Title: "…a 159-week prefecture-level sentinel surveillance study". Abstract: "analyzed weekly per-sentinel-facility COVID-19 case counts in all 47 prefectures of Japan". |
| | | (b) Provide in the abstract an informative and balanced summary of what was done and what was found | Structured abstract with Background, Methods, Results, Conclusions sections (348 words). |
| **Introduction** | | | |
| Background/ rationale | 2 | Explain the scientific background and rationale for the investigation being reported | Introduction §1 (post-Class-5 transition context); §2 (rationale for combining pseudo-entropy with Moran's *I*). |
| Objectives | 3 | State specific objectives, including any prespecified hypotheses | Introduction §3: three explicit objectives (i)–(iii). Hypotheses on metapopulation framework explicitly stated. The data- |

| Section / Topic | Item # | Recommendation | Page / Section in our manuscript |
|---|---|---|---|
| | | | driven commitment for identifying any structural change is explicitly noted as a methodological priority. |
| **Methods** | | | |
| Study design | 4 | Present key elements of study design early in the paper | Methods §"Study design": explicit declaration as ecological time-series study; STROBE compliance. |
| Setting | 5 | Describe the setting, locations, and relevant dates | Methods §"Setting and data sources": Japan, all 47 prefectures, 24 April 2023 – 7 May 2026 (159 weeks). |
| Participants | 6 | (a) Cohort study: give eligibility criteria, sources, methods of selection. (b) Case-control: give eligibility criteria. (c) Cross-sectional: give eligibility criteria, sources, methods of selection | (c) — Methods §"Setting and data sources": All 47 prefectures included; weekly per-sentinel-facility report rates extracted from MHLW open data. No selection or exclusion. |
| Variables | 7 | Clearly define all outcomes, exposures, predictors, potential confounders, and effect modifiers | Methods §§"Outcome measures": pseudo-entropy $S$, KL divergence regional decomposition, global, local, and time-lagged Moran's $I$, the spatial-persistence ratio $I(\tau)/I(0)$, all defined formally with equations. |
| Data sources / measurement | 8 | For each variable of interest, give sources | Methods §"Setting and data sources": |

| Section / Topic | Item # | Recommendation | Page / Section in our manuscript |
|---|---|---|---|
| | | of data and details of methods of assessment. Describe comparability of assessment methods if there is more than one group | MHLW sentinel surveillance system. URL provided. Per-sentinel-facility report rate as defined by Japan's Class-5 sentinel system. Sensitivity to the 7 April 2025 surveillance reform is reported in §"Sensitivity analyses" and Supplementary Materials. |
| Bias | 9 | Describe any efforts to address potential sources of bias | Methods §"Sensitivity analyses": five pre-specified sensitivity analyses including seven alternative spatial-weights matrices, harmonic-order choice $K \in \{1, \ldots, 6\}$ with AR(1) errors, leave-one-prefecture-out analysis for K = 2 vs K = 3 on Moran's *I*, surveillance-reform sensitivity, and Okinawa exclusion (46-prefecture re-analysis). Discussion §"Strengths and limitations" enumerates limitations including ecological-fallacy concerns and proxy validity. |
| Study size | 10 | Explain how the study size was arrived at | Methods §"Setting and data sources": 47 prefectures × 159 |

| Section / Topic | Item # | Recommendation | Page / Section in our manuscript |
|---|---|---|---|
| | | | weeks (full available post-Class-5 sentinel-surveillance record at time of analysis). |
| Quantitative variables | 11 | Explain how quantitative variables were handled in the analyses. If applicable, describe which groupings were chosen and why | Methods §§“Outcome measures” and “Change-point detection”: all formulas explicit. The pre-transition (n = 135 weeks) versus post-transition (n = 24 weeks) split was identified data-driven via the two-stage change-point detection procedure described in Methods §“Change-point detection”, not pre-specified. |
| Statistical methods | 12 | (a) Describe all statistical methods, including those used to control for confounding (b) Describe any methods used to examine subgroups and interactions (c) Explain how missing data were addressed (d) Cohort/cross-sectional: explain methods. (e) Describe any sensitivity analyses | Methods §§“Harmonic regression with autoregressive errors”, “Change-point detection”, “Statistical analysis”, and “Sensitivity analyses”: truncated Fourier regression with AR(1) errors estimated by Cochrane–Orcutt; data-driven change-point detection (PELT, CUSUM, Bai–Perron sup-F applied in a region restricted by the empirically established entropy ceiling); 9,999-permutation Moran’s *I* significance; |

| Section / Topic | Item # | Recommendation | Page / Section in our manuscript |
|---|---|---|---|
| | | | moving block bootstrap as principal inferential statistic for all between-period comparisons. The conventional Welch *t*-test was not used because of within-group serial dependence. (c) No missing weeks. (e) Five pre-specified sensitivity analyses described above. |
| **Results** | | | |
| Participants | 13 | (a) Report numbers at each stage of study. (b) Give reasons for non-participation. (c) Consider use of a flow diagram | (a) 47 prefectures × 159 weeks = 7,473 prefecture-week observations, all included. (b) None — public aggregate data, no individual participants. |
| Descriptive data | 14 | (a) Give characteristics of study participants and information on exposures and potential confounders (b) Indicate number of participants with missing data for each variable of interest. (c) Cohort: summarize follow-up time | Results §"Sample characteristics" (descriptive statistics for *S* and Moran's *I* across the full 159-week period); Supplementary Table S4 (week-by-week post-transition statistics). |
| Outcome data | 15 | Cohort: numbers of outcome events or summary measures over time. Case-control: numbers in | Results §§"Sample characteristics" through "Spatial persistence of clusters": numerical |

| Section / Topic | Item # | Recommendation | Page / Section in our manuscript |
|---|---|---|---|
| | | each exposure category. Cross-sectional: report numbers of outcome events or summary measures | results for *S(t)*, *I(t)*, KL contributions, lagged Moran's *I*, harmonic fits, regional shares, all reported. |
| Main results | 16 | (a) Give unadjusted estimates and, if applicable, confounder-adjusted estimates and their precision. (b) Report category boundaries when continuous variables were categorized. (c) If relevant, consider translating estimates of relative risk into absolute risk for a meaningful time period | Results §§"Wave-amplitude-invariant entropy ceiling", "Data-driven identification of a structural break in late November 2025", "The post-transition departure", and "Spatial persistence of clusters": all effect sizes reported with 95% bootstrap confidence intervals. The principal finding (spatial-persistence ratio shift of +0.62, 95% bootstrap CI +0.32 to +0.90, moving block bootstrap $p < 0.0001$) is reported as a between-period mean difference rather than as a ratio because the pre-transition mean is small relative to its standard deviation. The data-driven change-point at the week of 27 November 2025 is reported with multi-method consensus (PELT, CUSUM, Bai–Perron sup-F all within a 2-week window). (c) |

| Section / Topic | Item # | Recommendation | Page / Section in our manuscript |
|---|---|---|---|
| | | | N/A — ecological design. |
| Other analyses | 17 | Report other analyses done — e.g., analyses of subgroups and interactions, and sensitivity analyses | Results §“Sensitivity analyses”: summary of the five pre-specified sensitivity analyses. Supplementary Materials §§S2–S8 expand on these in full. Global change-point detection on the unrestricted 159-week series (S8) is reported as a baseline comparison to the principal restricted analysis. |
| **Discussion** | | | |
| Key results | 18 | Summarize key results with reference to study objectives | Discussion §“Principal findings”: the three pre-specified objectives are addressed in turn, with explicit reference to the data-driven identification of the late-November 2025 change-point. |
| Limitations | 19 | Discuss limitations of the study, taking into account sources of potential bias or imprecision. Discuss both direction and magnitude of any potential bias | Discussion §“Strengths and limitations”: six explicit limitations including the 24-week post-transition observation window, prefecture-level aggregation, absence of direct mechanistic identification, the recent partial re-emergence of Okinawa case shares |

| Section / Topic | Item # | Recommendation | Page / Section in our manuscript |
|---|---|---|---|
| | | | (raising the possibility of a longer cycle rather than a stable new regime), the limited ability of change-point detection to identify latent mechanisms, and the n = 19 dataset version. |
| Interpretation | 20 | Give a cautious overall interpretation of results considering objectives, limitations, multiplicity of analyses, results from similar studies, and other relevant evidence | Discussion §§"Comparison with prior literature", "Mechanistic interpretation", "Implications for endemic transition theory": interpretation under metapopulation, broken-link, and endemic-transition frameworks. Emphasis on "candidate regime transition" rather than confirmed regime change. Comparison with Manz 2021, Tokumoto 2025, Belvis 2023 explicit. |
| Generalizability | 21 | Discuss the generalizability (external validity) of the study results | Discussion §"Implications for endemic transition theory" and Conclusions: the two-mode anti-phase framework potentially generalizes to influenza and other respiratory pathogens within the |

| Section / Topic | Item # | Recommendation | Page / Section in our manuscript |
|---|---|---|---|
| | | | same surveillance infrastructure; cautions against uncritical extrapolation to other countries with different sentinel-system designs. |
| **Other information** | | | |
| Funding | 22 | Give the source of funding and the role of the funders for the present study and, if applicable, for the original study on which the present article is based | Statements §"Funding": institutional support from The University of Osaka (RCNP, CiDER, CAMaD); no direct external research funding for this specific work. |

***Note:*** *This study is an ecological time-series analysis of fully aggregated, publicly available prefecture-level surveillance data. It does not involve individual-level participants, exposures, or outcomes. Items concerning individual-level data collection, missingness, or selection bias are addressed at the prefecture-week level where applicable. The completed checklist follows the STROBE statement adapted for cross-sectional ecological studies.*
